# Optimization and performance analytics of global aircraft-based wastewater surveillance networks

**DOI:** 10.1101/2024.08.02.24311418

**Authors:** Guillaume St-Onge, Jessica T. Davis, Laurent Hébert-Dufresne, Antoine Allard, Alessandra Urbinati, Samuel V. Scarpino, Matteo Chinazzi, Alessandro Vespignani

## Abstract

Aircraft wastewater surveillance has been proposed as a novel approach to monitor the global spread of pathogens. Here we develop a computational framework to provide actionable information for designing and estimating the effectiveness of global aircraft-based wastewater surveillance networks (WWSNs). We study respiratory diseases of varying transmission potentials and find that networks of 10 to 20 strategically placed wastewater sentinel sites can provide timely situational awareness and function effectively as an early warning system. The model identifies potential blind spots and suggests optimization strategies to increase WWSNs effectiveness while minimizing resource use. Our findings highlight that increasing the number of sentinel sites beyond a critical threshold does not proportionately improve WWSNs capabilities, stressing the importance of resource optimization. We show through retrospective analyses that WWSNs can significantly shorten the detection time for emerging pathogens. The presented approach offers a realistic analytic framework for the analysis of WWSNs at airports.

---

Recent health crises have highlighted the dual role of airports both in spreading infectious diseases globally and simultaneously acting as convenient frontlines for detecting and monitoring emerging health threats [1–5]. In this context, aircraft-based wastewater surveillance is gaining increasing scientific and operational interest. Traditionally, wastewater surveillance has been used to monitor community prevalence of pathogens such as SARS-CoV-2 variants [6, 7], polio [8, 9], and influenza [10, 11]. Expanding wastewater surveillance at airports to create a global wastewater surveillance network (WWSN) has been recently proposed as a novel, early warning system against emerging pathogens [12–17]. Sampling aircraft wastewater provides a noninvasive method to monitor the spread of pathogens, but the creation of a global WWSN poses strategic and operational challenges. These challenges include efficient sample collection, logistics for genomic analysis, decisions such as what pathogens to test for, selecting optimal airports for surveillance, scaling the network, and addressing surveillance blind spots to balance effectiveness and cost [18]. While there have been studies on the feasibility of aircraft wastewater surveillance at several major airports [19–22], fully understanding the performance of a WWSN—in terms of its size, distributed locations, and operations—remains to be addressed.

Here, we use the Global Epidemic and Mobility Model (GLEAM) [23–25] to study the performance of global WWSNs, providing valuable insights into how pathogens spread and are detected within these networks. GLEAM is a stochastic, spatial, age-structured metapopulation model. It divides the global population into over 3,200 subpopulations across approximately 200 countries and territories, all interconnected by air travel and commuting networks. The air travel component of the model includes data on flight segments and origin-destination information for more than 4,600 airports, obtained from the Official Aviation Guide (OAG) database (see Methods and Sec. 1 in the Supplementary Information, SI). The mobility framework is coupled with an epidemic compartmental model tracking individuals within various disease stages (ex. Susceptible, Latent, Infectious, etc.) and their movement across subpopulations at a global level. GLEAM has been successfully applied to model global health threats including pandemic influenza, Ebola, Zika, and SARS-CoV-2 [26–28]. To simulate a surveillance system within GLEAM, we create a global WWSN that consists of multiple surveillance sites—called *sentinels*. We assume each sentinel airport will test the wastewater from a certain fraction of arriving international flights per day.

Given any initial conditions for an outbreak, the model generates stochastic realizations of the global epidemic spread. Simulated data include international and domestic infection importations, incidence of infections, and individual-level detection at sentinel sites with a daily resolution. The early growth phase of the modeled epidemics can be mapped onto a multitype branching process allowing for the computationally efficient derivation in terms of probability generating functions (PGFs) of several key analytics that quantify the efficiency of the WWSN. These include the time to first detection of an emerging pathogen and, from measured detections at sentinels, the identification of the source location, the estimation of the reproduction number, and the timing of the outbreak’s onset. These metrics provide a general framework for assessing the WWSN’s effectiveness in real-time for surveillance and public health response.

## Results

We start our analysis by considering a baseline Wastewater Surveillance Network (WWSN) with 20 sentinel sites. To achieve sufficient regional coverage, we selected the three busiest international airports from each of the six World Health Organization regions and added two additional sites in South America and Oceania. The locations are shown by airport markers in Fig. 1 and reported in Table S4 in the SI.

**Figure 1:**
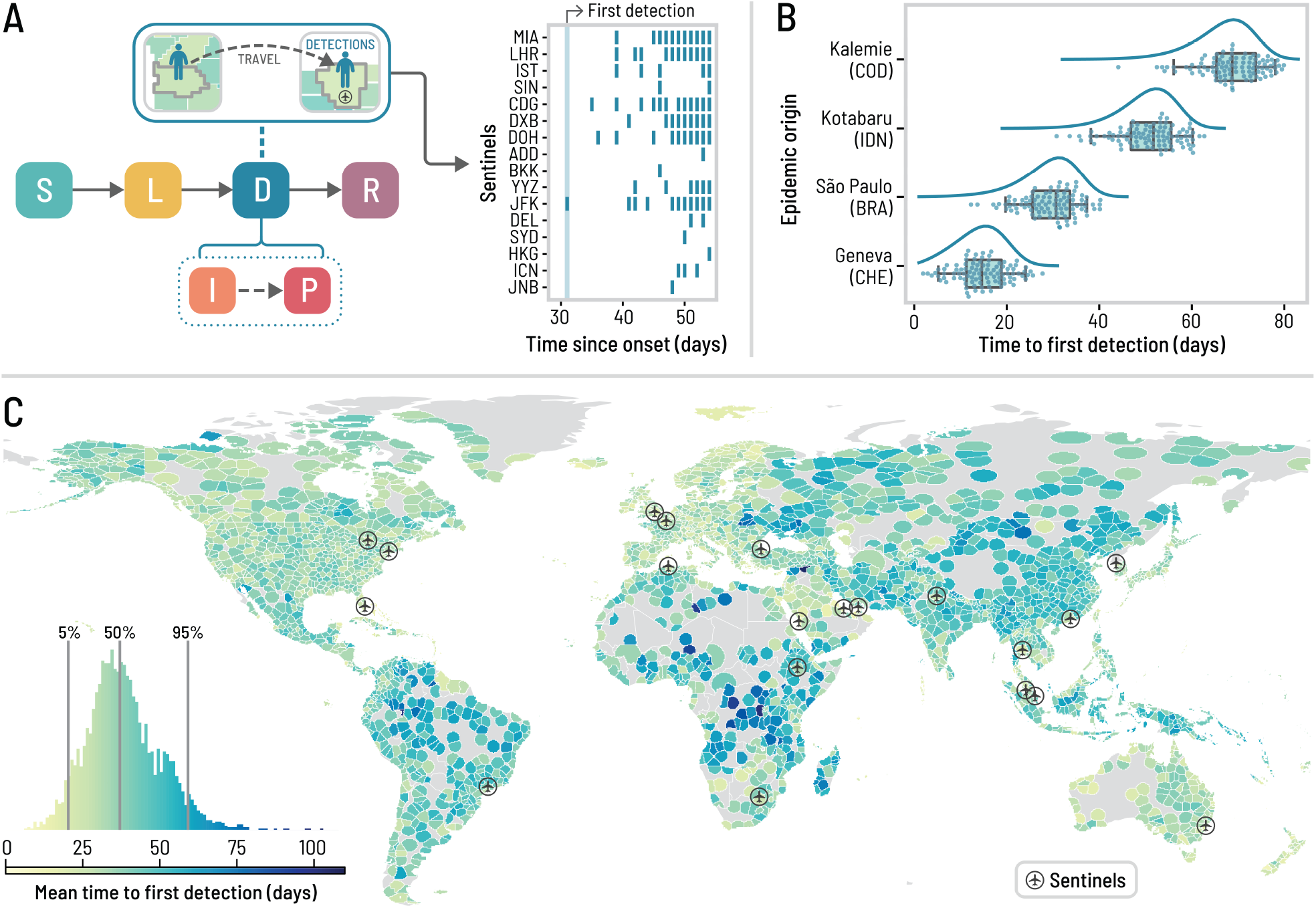
Time to detect a novel pathogen with a global surveillance network at airports. The surveillance system corresponds to a baseline network of 20 sentinel airports, chosen based on their high volume of international passengers and favoring geographical diversity (see Table S4 in the SI). We use an average basic reproduction number ⟨ℛ_0_⟩ = 2 at the source, a generation time of 4 days, and a post-infectious period of 10 days, resulting in a detectable period of ∼12.7 days. For detectable individuals, the probability of detection on an international flight incoming to a sentinel is 16%. (A) Schematic of the Susceptible–Latent–Detectable–Recovered (SLDR) compartmental model and example of binary detection time series at sentinel airports generated by GLEAM, utilizing São Paulo as the origin. Only sentinels (identified by their IATA code) with at least one detection are shown. (B) Time to first detection by the sentinel system for four different origins (with the ISO 3166-1 alpha-3 codes of the origin countries in parentheses). Each dot is a GLEAM simulation and the box plot summarizes the results (*n* = 100 for each origin). The center line of the box plot indicates the median, the box covers the interquartile range and the whiskers cover the 90% central prediction interval. The curves correspond to analytical distributions from the PGF methodology. (C) Mean time to first detection by the surveillance network, considering each subpopulation as the potential origin for an epidemic. Each subpopulation is colored according to the mean time to first detection if it were the origin of an outbreak. The histogram in the lower left corner compiles the results from 3244 subpopulations.

The efficiency of the WWSN is related to the intrinsic characteristics of a pathogen as well as its detectability. To test the effectiveness of the WWSN we assume a SARS-CoV-2-like respiratory infection is spreading with a detectable period in wastewater similar to what is reported in Refs. [6, 29, 30]. We map an individual’s disease history to a Susceptible–Latent–Detectable–Recovered (SLDR) compartmental structure, as shown in Fig. 1A. Susceptible (S) individuals can get infected through exposure to infectious individuals. Latent (L) individuals have been exposed but are not yet transmitting the pathogen and remain undetectable in the wastewater. Detectable (D) individuals include both infectious (I) individuals who can transmit the pathogen and post-infectious (P) individuals who no longer infect others but are still detectable through wastewater. Finally, recovered (R) individuals are no longer detectable and cannot be reinfected (See Methods for details and key time-to-event intervals).

Each traveling detectable individual arriving at a sentinel on an international flight is detected with probability *p*_det_. The detection rate *p*_det_ combines the fraction of sampled aircraft, the probability an individual uses the lavatory during a flight, and the probability a detectable individual is shedding enough virus to lead to a detection. Current detectability estimates for SARS-CoV-2 in aircraft wastewater vary considerably [14, 16, 21], therefore, our analysis varies the detection probabilities, *p*_det_, from 4 to 32% (see Methods for a detailed discussion). While sampling individual aircraft independently is optimal for detection accuracy, it may be more cost-effective to test combined wastewater from multiple aircraft at a consolidation point such as the airport triturator. Consequently, we assume that through pooled sampling, multiple detectable individuals traveling through the same sentinel on the same day cannot lead to more than *one detection*, leading to binary detection time series as illustrated in Fig. 1A. It is worth highlighting that most of these assumptions can be relaxed in order to use different detection schemes, cadence, and sentinel site locations.

### Baseline WWSN performance

A key metric for evaluating the effectiveness of a WWSN is the *time to first detection t*_fd_ of an emerging pathogen. This metric is defined as the number of days from the onset of an outbreak until the first detection at any sentinel. In our analysis, we seed an epidemic in a single subpopulation with a small, initial cluster of 10 latent and 10 infectious individuals. The time to first detection inherently depends on the WWSN configuration, the outbreak’s origin, the pathogen characteristics, and the operational detection rate. At the same time, there are also fluctuations stemming from the stochastic nature of each outbreak and the individuals’ traveling and detection events.

In Fig. 1B, we show the full probability distribution *P* (*t*_fd_) for the time to first detection for four different origins: Geneva (Switzerland), São Paulo (Brazil), Kotabaru (Indonesia), and Kalemie (Democratic Republic of the Congo). In this figure, we assume that the detection rate in the WWSN is *p*_det_ = 16% and is uniform across all 20 sentinels. We see that the time to first detection can vary significantly, with a mean of 14.2 (90% PI, 4–22) days for Geneva, to 66.5 (90% PI, 53–76) days for Kalemie, where PI refers to the central prediction interval (5th and 95th percentiles for the 90% PI). To obtain a global picture of the WWSN performance for different epidemic origin locations, we calculate the mean time to first detection, *T*_fd_, for each potential source across all of the 3,200+ subpopulations in our model (see Fig. 1C). A notable aspect is the significant spatial variability of *T*_fd_ based on the epidemic’s origin. For certain locations in Central Africa, *T*_fd_ is of the order of 100 days, while for many places in Europe, 15-25 days is more typical. This heterogeneity is further highlighted in Fig. 2 at the level of continents. While Fig. 2 shows that epidemics emerging from some continents take more time than others to be detected by a global WWSN, we note an important heterogeneity within continents as well. For instance, in Africa, the 90% PI of the *T*_fd_ ranges from 23 to 71 days. Even looking down at the level of statistical subregions as defined by the United Nations geoscheme in Fig. S5 of the SI, we still find very broad distributions of *T*_fd_ for all subregions. Middle Africa for instance is very dispersed, with a 90% PI ranging from 28.2 to 84.5 days. This means that within all regions, and at various scales, there are potential *blind spots* for a global WWSN: locations that if they are the source of an epidemic, will take a very long time to lead to a detection.

**Figure 2:**
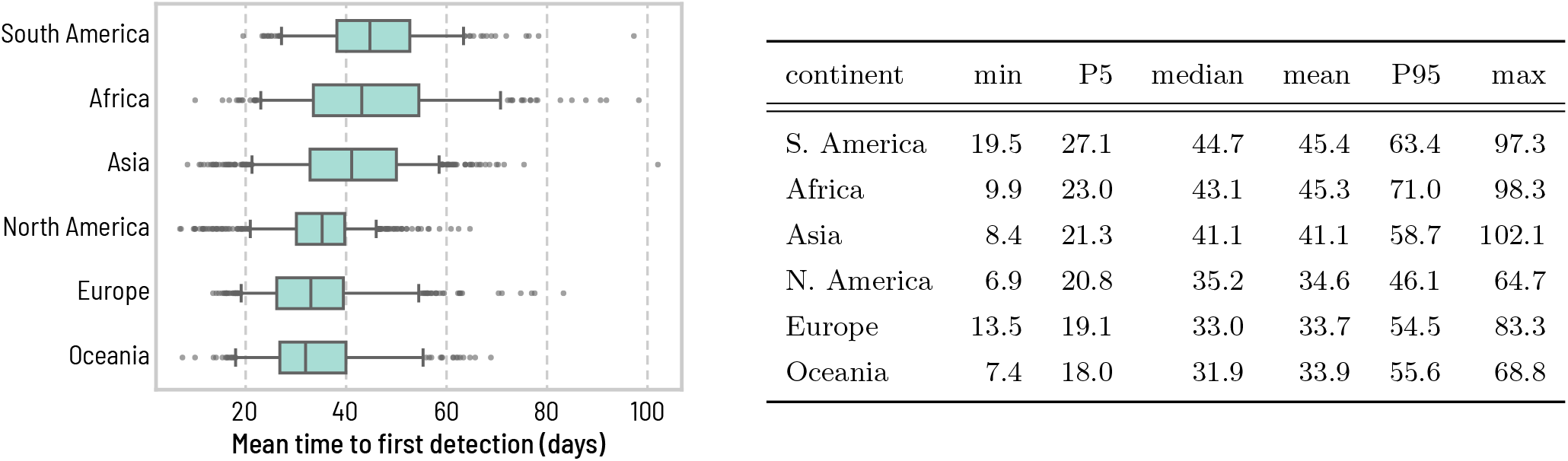
Heterogeneity of the time to first detection within geographical regions. We aggregate the mean time to first detection *T*_fd_ obtained in Fig. 1 over continents (S. America, *n* = 297; Africa *n* = 338; Asia *n* = 867; N. America *n* = 854; Europe *n* = 596; Oceania *n* = 292). The center line of the box plot indicates the median, the box covers the interquartile range, the whiskers cover the 90% central prediction interval (P5–P95), and black dots correspond to outliers outside this interval. Numerical values for some of the statistics of the mean time to first detection are reported in the table on the right.

These blind spots within the WWSN can be attributed in part to the per capita volume of travel. In the SI we show a strong indirect correlation between the per capita volume of travel and the mean time to first detection (Fig. S7). However, this is not the only factor contributing to blind spots: detections at sentinel sites are not solely the result of direct importations from the outbreak’s origin, but sometimes rely on importations from secondary outbreak locations experiencing community transmission. This indirect path to reach a sentinel further increases the detection time.

### Effects of pathogen characteristics on the WWSN

It is important to note that the natural history of a disease, particularly its key characteristic times and reproductive number, significantly impacts the detection time (*T*_fd_). In Fig. 3A, we show how the global distribution of *T*_fd_, aggregated over all locations, changes as the reproduction number ℛ_0_, the generation time *T*_gen_, and the surveillance detection rates *p*_det_ are varied. A larger reproduction number and a shorter generation time lead to shorter *T*_fd_, and vice-versa; the smaller the probability of detection the longer *T*_fd_, although with limited impact. This can be explained by the exponential growth of epidemics in their early stages. The WWSN will typically start detecting cases when there is a sufficient number of detectable individuals *D* traveling through the WWSN. This number is approximately 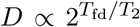, where *T*_2_ is the *doubling time* of the epidemic (here measured in days). Adjusting the basic reproduction number, ℛ_0_, or the generation time, *T*_gen_, significantly affects the time to first detection due to alterations in *T*_2_. Conversely, changes in the detection probability *p*_det_ do not similarly impact the timing. Indeed, a two-fold reduction in *p*_det_ implies a two-fold increase in *D* before detection. However, this increase in *D* happens in the span of a single doubling time, *T*_2_. The exponential growth also implies that the ratio *T*_fd_*/T*_2_ should be approximately constant as we vary the doubling time of the epidemic. More precisely, as shown in Fig. 3B, the complete invariant quantity reads as

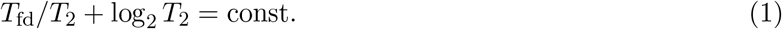

**Figure 3:**
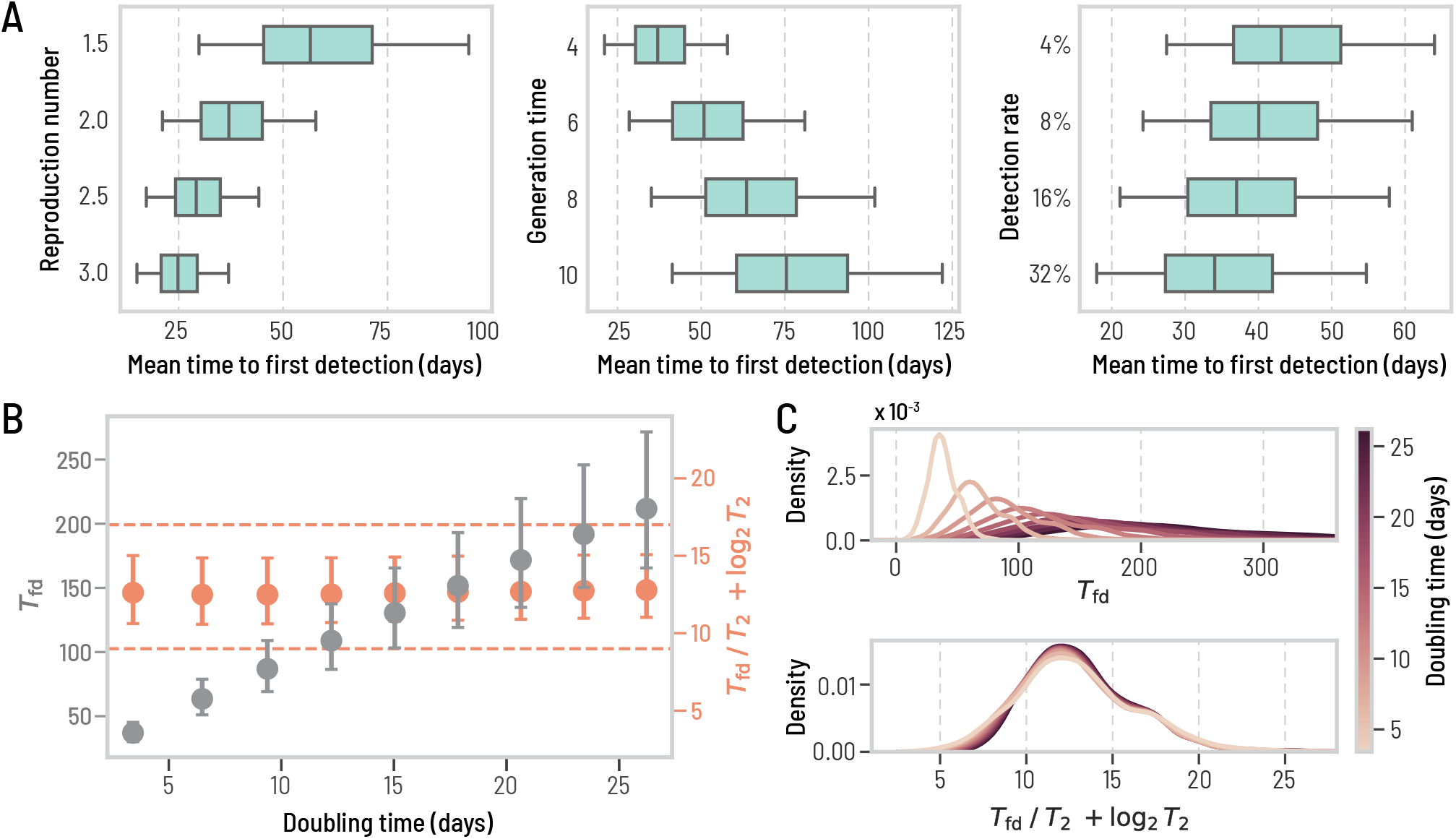
Changing the transmission dynamics predictably affects the time to first detection. We use the same baseline WWSN as in Fig. 1 and the same detectable period. Unless specified, we keep an average reproduction number of 2, a generation time of 4 days, and a 16% detection rate at sentinels. All prediction intervals are obtained with *n* = 3244 subpopulations. (A) *T*_fd_ from all origins, with varying reproduction number, generation time, and detection rate. The center line of the box plot indicates the median, the box covers the interquartile range and the whiskers cover the 90% central prediction interval; the outliers outside the interval are not shown. (B-C) We vary the generation time between 4 and 36 days, resulting in doubling times between 3.4 and 26.2 days. (B) *T*_fd_ and *T*_fd_*/T*_2_ + log_2_ *T*_2_ as a function of the doubling time. Circles indicate the median and the error bars cover the interquartile range. The dashed lines are there to guide the eyes. (C) Distributions of *T*_fd_ and *T*_fd_*/T*_2_ + log_2_ *T*_2_ over all origins for different doubling times. We use kernel density estimates (KDE) for the distributions to improve the visualization.

The correction term log_2_ *T*_2_ is necessary to account for the stochastic nature of the detection process [31] (see SI, Sec. 2). In Fig. 3C, we also show how the distributions of mean time to first detection collapse onto one another when considering the invariant quantity in Eq. (1). In practical terms, altering the disease characteristics effectively results in a linear transformation of *T*_fd_ across all locations (see also Fig. S9 in the SI). Therefore, focusing on a specific parametrization does not result in any loss of generality of the results, allowing for consistent and generalizable analyses. Other aspects of disease transmission affecting *T*_fd_—overdispersion of the secondary-infection distribution, length of the detectable period, and seasonal change in the air-travel network—have a very limited impact (see Table S3 in the SI).

### Scaling and optimization of WWSNs

Key features like the number of sentinel airports and their geographical placement are crucial for optimizing the effectiveness of the WWSN. The strategic optimization of these features represents a classic resource-constrained optimization problem. To systematically assess the network’s efficiency, we define *T*_fd_(𝒮, *l*) as the mean time to first detection for a WWSN configuration. Here, 𝒮 denotes the set of sentinel sites, and *l* indicates the subpopulation at the epidemic’s origin. We can then average this metric over multiple origins *l* by weighing each location according to a prior distribution *P* (*l*) for the occurrence of an outbreak, resulting in the (average) mean time to first detection

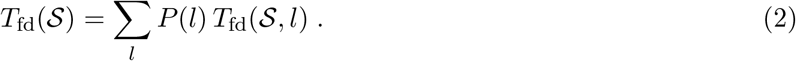

While *T*_fd_(𝒮) is a well-defined indicator of performance, its value is sensitive to variation of the disease transmission characteristics (see Eq. (1)). To provide a more informative measure of network efficiency, we compare *T*_fd_(𝒮) with *T*_fd_(𝒞), where the latter represents the average mean time to first detection for a hypothetical complete WWSN that includes all international airports globally. This comparison helps us quantify the relative performance of a specific sentinel configuration 𝒮. We define the excess time to first detection for the sentinel system 𝒮 using the following formula:

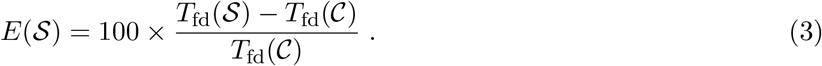

This metric represents the additional percentage of time it takes for the system 𝒮 to achieve its first detection compared to the complete network.

We use three different strategies to define the geographic distribution of the sentinel network: (1) ranking airports based on their international inbound passenger *volume* [15], (2) ranking airports by their *entropy* in traffic flows—a measure of diversity that favors airports offering wide geographical connectivity, and (3) using a *greedy* optimization strategy that minimizes the mean time to first detection (see Methods). In Fig. 4A, we show the excess time to first detection for the three different strategies considered, assuming a homogeneous prior for the source of an epidemic, irrespective of the area or population size (i.e., *P* (*l*) = const. for all *l*). While the greedy approach systematically provides the smallest excess time, all three strategies have similar performances, despite different network configurations (see Table S4). The radar chart also suggests that the greedy strategy provides a relatively balanced geographical surveillance when compared to the complete WWSN. The most striking result of the optimization analysis, however, resides in the clear case of diminishing returns when increasing the number of sentinels. Our findings show that a network with 20 sentinels would only take approximately 20% longer to achieve a first detection compared to a system that includes thousands of airports. Furthermore, doubling the number of sentinels from this number results in marginal gains, improving detection time by less than 10%. This result indicates a highly cost-effective trade-off between the efficiency of the Wastewater Surveillance Network (WWSN) and the resources allocated to it. With just a small number of sentinels, the network achieves nearly the full efficiency of a system comprising thousands of airports.

**Figure 4:**
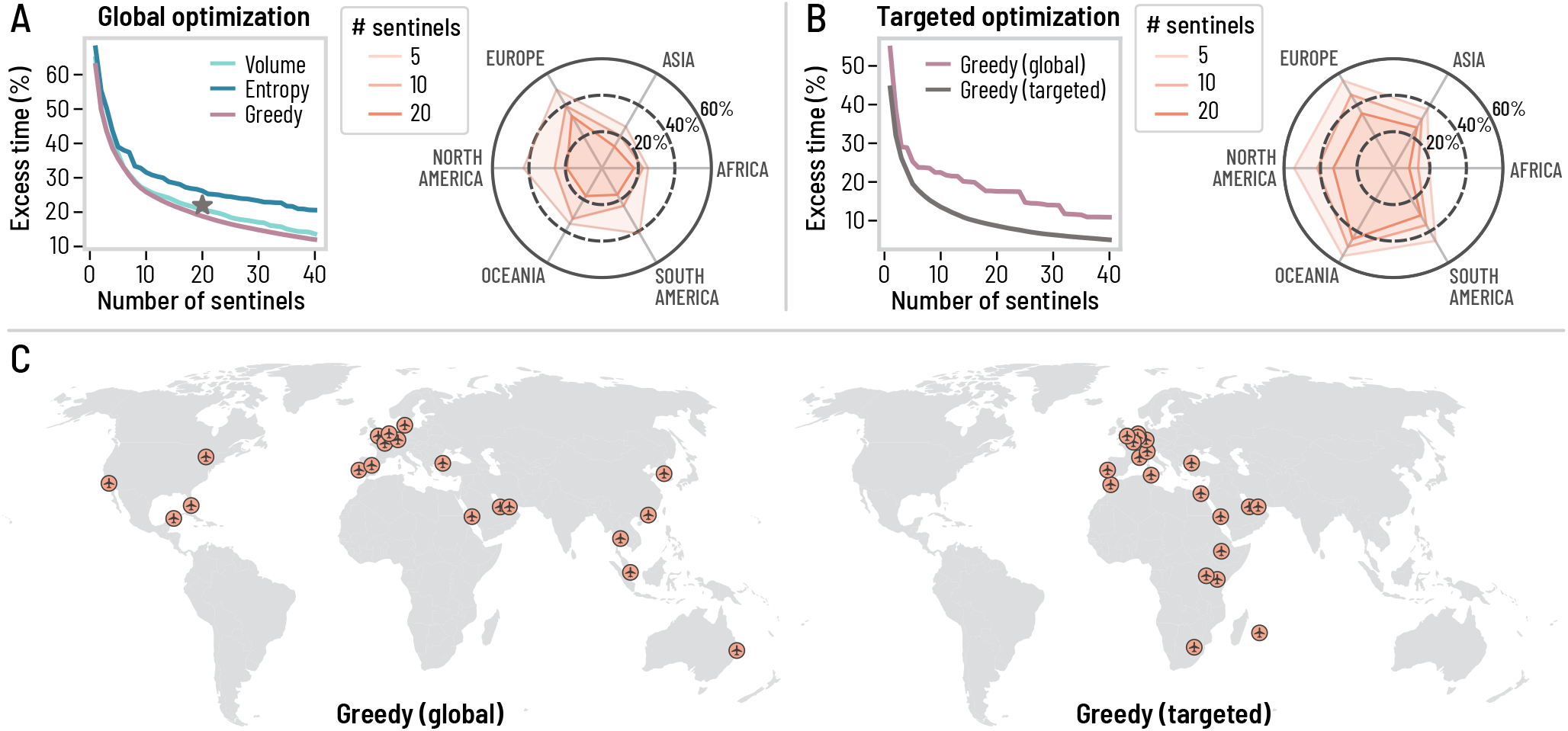
Scaling and optimization of a global surveillance network at airports. We use the same disease parametrization as in Fig. 1, but we use different strategies to choose the sentinel airports and we vary their number. We evaluate *T*_fd_ and compute the excess time to first detection compared to the complete WWSN. (A) Global optimization: we assume all subpopulations are equiprobable for the origin of an epidemic. The star corresponds to the excess time for the baseline network. (B) Targeted optimization: the greedy strategy is tuned to minimize the excess time for epidemics originating from Africa. (A-B) The radar charts illustrate the excess time to first detection using the greedy strategy (global and targeted), assuming the origin is within a specific continent. A balanced strategy will minimize the excess time equally for all geographical regions, while a smaller excess time for a specific region indicates a bias, the intended feature of a targeted optimization procedure. (C) Spatial distribution of the 20 first sentinels for the global and targeted greedy optimization strategies.

Some diseases are only endemic in certain parts of the world or have clear seasonal patterns. Besides, we have shown that *T*_fd_ is higher for some geographical areas than others (see Fig. 2). For these reasons, it might be relevant to tailor the WWSN to specific geographical areas by biasing the optimization procedure to improve the detection capabilities for specific regions or subpopulations. The greedy optimization approach can be easily adapted to do so by tuning the prior function *P* (*l*). For instance, if we aim to minimize the excess time to first detection for epidemics originating in Africa, we can set *P* (*l*) = const. if *l* is in Africa, otherwise *P* (*l*) = 0. In Fig. 4B, we present the excess time to first detection and compare the conventional global greedy optimization method with our targeted greedy optimization strategy. The radar chart in Fig. 4B illustrates the bias introduced by the targeted optimization strategy, particularly showing a skewed coverage favoring the African continent at the expense of performance in other regions. The maps in Fig. 4C show how sentinel placement changes markedly when the optimization process targets a specific geographical area; focusing on the African continent results in a higher concentration of sentinels in Africa and Europe, aligning with traffic flow considerations. These results open the paths to optimize the surveillance system dynamically, adapting the WWSN to evolving knowledge on the epidemic and its geographical dispersion.

### Situational awareness with WWSNs

WWSNs can be used to provide evolving situational awareness on emerging infectious diseses threats. To illustrate the potential use of WWSNs to gather epidemiological information, we explore the emergence of the SARS-CoV-2 Alpha variant (B.1.1.7) in Fall 2020 (see SI Sec. 3 for details) [32–34]. More precisely we consider a hypothetical scenario where the baseline WWSN illustrated in Fig. 1 is assumed to be operational. The study uses air-travel data from September to November 2020 and in Fig. 5A, we present probable distributions for the time to first detection of the Alpha variant. Our findings indicate that, even with the 4% detection rate, the variant would likely have been detected as early as November. Specifically, the median time to first detection is projected to be November 13, with a 90% PI ranging from October 15 to December 1. With a 16% detection rate, our analysis suggests that the first detections of the Alpha variant would likely have occurred by late October, with a median detection date of October 29, 2020, and a 90% PI from October 2 to November 16. Given that the Alpha variant was first reported by the United Kingdom government on December 14, 2020, this analysis shows the potential of a global WWSN to serve as an effective early warning system.

**Figure 5:**
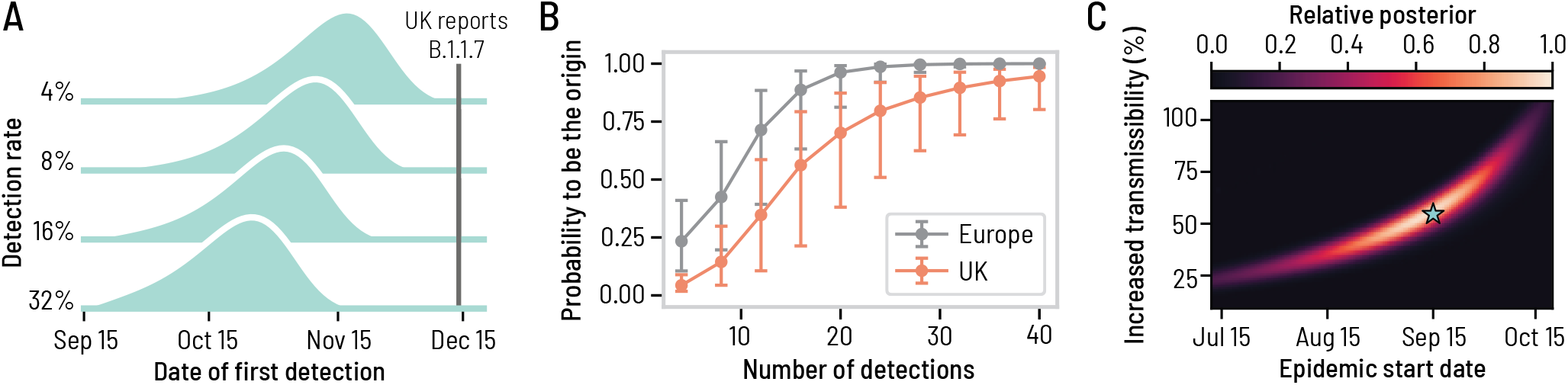
A global WWSN would provide an early warning system for international spreading and timely inferential capabilities. We consider a counterfactual scenario of the emergence of the SARS-CoV-2 Alpha variant where a global WWSN would have been available. We use the baseline surveillance system illustrated in Fig. 1. We considered an effective reproduction number of 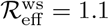 for the wild strain SARS-CoV-2 virus, and an increased transmissibility of 55% for the Alpha variant, resulting in an effective reproduction number of 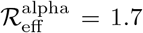. We also assumed a generation time of 6.5 days, and a post-infectious period of 10 days. We assume an initial cluster of 20 infectious and 20 latent individuals in the London and South East England region on September 15, 2020. (A) Distributions for the time to first detection with varying detection rates. (B)-(C) Inference experiment using data generated by the mechanistic GLEAM model, assuming a 16% detection rate. (B) Geolocalisation of the source as more detections cumulate. We compute the posterior distribution for the origin of the epidemic based on the detection counts at each sentinel. The markers indicate the median posterior value and the error bars cover the interquartile range obtained from 1250 detection time series. (C) Joint posterior distribution on the increased transmissibility of the Alpha variant and the epidemic start date, averaged over 125 possible instances of the detection time series. Gouraud interpolation is used to improve the visualization. The blue star indicates the ground truth values for the simulation experiment, corresponding to September 15 and an increased transmissibility of 55%.

Alongside tracking the initial international spread, the WWSN can also deliver timely information on the origin of an outbreak and help in understanding its growth dynamics. In Fig. 5(B), we show the probability that the WWSN correctly identifies the continent and country of origin as multiple detections accumulate in the system. This is achieved by calculating the posterior distribution *P* (*l*|***d***) for each subpopulation *l* to be the origin of an epidemic based on the cumulative number of detections at each sentinel ***d*** = (*d*_*ν*_)_*ν∈S*_ (see SI Sec. 3). Our analysis indicates that the source country could have been accurately identified after approximately 20 detections, which would likely have occurred by December 5th in over 50% of the model’s realizations with a 16% detection rate. In practice, a more efficient adaptive localization strategy could be employed. This would involve targeted sampling from individual aircraft in regions suspected to be the epicenter of the outbreak, thereby enabling more precise and timely identification of the source.

Multiple detection events can also be utilized to estimate key epidemic parameters, such as the growth rate, the onset time, and, given some knowledge of the generation time of the contagion process, the reproduction number of the epidemic. Here, we focus on the inference of the epidemic start date and the increased transmissibility of the Alpha variant compared to the original SARS-CoV-2 strain. As of December 14, 2020, the time at which the United Kingdom first reported the Alpha variant, the time series of detection events across the sentinel sites would yield the joint posterior distribution of the inferred epidemic start date (90% CI, July 29–Oct. 12) and increased transmissibility (90% CI, 25–91%) reported in Fig. 5C. The high posterior density region also matches closely the value of the simulation experiment, i.e., a start date of September 15 and a 55% increased transmissibility. The detailed inference procedure and individual posterior distributions for different time series are reported in Sec. 3 of the SI. Additional evidence of the timely situational awareness capacities provided by a global WWSN is presented in the SI Sec. 3 with the hypothetical scenario where a global WWSN would have been operational at the time of the emergence of SARS-CoV-2 in Wuhan.

## Discussion

The modeling framework we have presented offers a systematic analysis of global wastewater surveillance networks (WWSNs) at airports, their potential to characterize the international spread of diseases, and their capability to provide timely insights about an unfolding epidemic. Our findings demonstrate the significant role of WWSNs in shortening the time to first detection of pathogens, overcoming some of the challenges faced with standard symptoms-based passenger screening across regions [35]. It is important to emphasize that gaining even a few additional days of situational awareness regarding the introduction of a pathogen can be crucial in effectively controlling an outbreak. Moreover, in scenarios like the one we analyzed for the B.1.1.7 SARS-CoV-2 variant, the sentinel system could also be utilized for retrospective data assessment. This analysis can help establish the date of pathogen introduction and the potential geographical spread, enabling the optimization of travel restrictions and border screening policies. While these measures are costly, they are frequently implemented too narrowly or too late, diminishing their effectiveness. Wastewater Surveillance Networks (WWSNs) are ideally positioned to provide timely and precise surveillance data, enabling more effective public health responses. Finally, our framework allows the identification of the potential blind spots of each WWSN, paving the way to the integration of complementary surveillance methods, such as environmental wastewater monitoring in communities to cover gaps and enhance the network’s overall efficacy [36].

The strategies and optimization experiments conducted here were designed to demonstrate the capabilities of a WWSN rather than tailor them to a specific disease or outbreak. Future studies should therefore broaden the application of the modeling framework by integrating knowledge and experience developed for the surveillance of specific pathogens like arboviruses and influenza [37, 38] for instance. Additionally, incorporating factors influencing zoonotic spillovers—shaped by socioeconomic, environmental, and ecological dynamics—will enhance our understanding of emerging diseases and our predictive capabilities [39–42].

Future modeling work should also integrate the specific logistic capabilities of the WWSN with a thorough exploration of operational implementation such as rotating testing schedules and cadence across different sentinel sites in order to navigate logistical constraints. In other words, the modeling framework should be implemented alongside the actual deployment of the surveillance system, considering the practicalities and challenges of on-site operations. One such practical consideration highlighted by our study is the implementation of targeted and adaptive strategies that would not only improve pathogen screening efficiency but also ensure resource-efficient network operation.

Like all modeling studies, our analysis contains assumptions and limitations that must be clearly identified. Our approach considers air travel as an independent process for each individual and thus neglects clusters and household travel. On a more technical side, the analytics developed in this study rely on a multitype branching process, which neglects saturation effects from finite population sizes. Although these effects are minor and do not alter our conclusions at the early stage of an outbreak, they should be carefully considered when analyzing the performance of the WWSN for the inference of incidence and prevalence of large epidemics or endemic situations. The modeling framework also does not consider false positive test results and the occurrence of positive tests caused by wastewater tanks not being cleaned in between flights [21]. While this should not impact substantially our analysis of the time to first detection, it could affect the situational awareness capabilities of the WWSN. Therefore, future analysis should incorporate the prior probability of a pathogen circulating in the statistical model, along with the test specificity and the possibility of wastewater tank contamination between flights. This will be important for decision-making, especially if rare but very high-consequence pathogens are detected.

Taking into account these limitations, our study offers critical insights for analyzing wastewater surveillance at airports, providing a robust computational platform for informed public health decision-making. The quantitative insight provided by our approach holds significant implications for a range of stakeholders in public health, policies, and global health security. Furthermore, although our study focuses on airport wastewater surveillance, the proposed modeling framework can be adapted to environmental monitoring and other travel-based surveillance methods, such as nasal swab testing, thereby providing a full computational framework for the analysis of genomic, travel-based disease surveillance systems.

## Methods

### The Global Epidemic and Mobility model

The Global Epidemic and Mobility model (GLEAM) is a computational platform used for modeling epidemic spread, combining stochastic elements and spatial data in an age-structured, metapopulation framework [23–25]. GLEAM divides the world into distinct geographic subpopulations using a Voronoi tessellation of the Earth’s surface, with each subpopulation centered around major transportation hubs such as airports. These subpopulations are detailed with high-resolution data about population demographics, age-specific contact patterns, health infrastructure, and other relevant attributes based on available data. GLEAM incorporates a human mobility layer into its modeling, using data from various sources, including the Official Aviation Guide (OAG) and IATA databases. This layer includes both short-range (e.g., commuting) and long-range (e.g., flights) mobility data, and creates a network of daily passenger flows between airports worldwide. The model uses a worldwide homogeneous standard for commuting and compensates for missing information with synthetic data based on the “gravity law” calibrated with real data [23, 25].

GLEAM tracks the number of individuals in each disease state for all subpopulations over time. It simulates travelers’ movements through the flight network, with air travel probabilities varying by age group. Finally, the disease dynamics and the detection process at airports within GLEAM are simulated using stochastic binomial chain processes. These processes rely on parameter values sourced from existing literature, defining the natural history of the infection being modeled. We refer to this implementation as the *mechanistic* GLEAM approach. See Sec. 1 of the SI for a more technical description of the model. All our analyses make use of a global air-travel network capturing the period of September 2022 to August 2023, except the case studies on the emergence of COVID-19 and the SARS-CoV-2 Alpha variant where we use data from December 2018 to February 2019^1^ and September to November 2020 respectively.

### Disease progression and transmission dynamics

To model the disease transmission within the subpopulations and the detections at airports following air travel, we make use of a standard compartmentalization scheme for the disease progression. Each individual, at any time point, is assigned to a compartment corresponding to their particular disease-related state. An individual who gets infected will go through the following sequence of states: susceptible (S; pre-exposure), latent (L; exposed, but do not yet transmit the infectious pathogen), infectious (I; can transmit the disease), post-infectious (P; no longer infectious), and recovered (R). In our model, we assume that only infectious and post-infectious can be detected through wastewater, which we regroup under the *detectable* (D) state. The inclusion of the post-infectious state in our model is necessary because diseases such as COVID-19 remain detectable in wastewater well beyond the active infectious period. [6, 29, 30].

The contagion dynamics in each location is strongly influenced by the basic reproduction number ℛ_0_— the average number of secondary infections caused by an infectious individual in a fully healthy population. In our model, the reproduction number ℛ_0_ varies slightly from one subpopulation to another, since it is proportional to the largest eigenvalue of the age-structured contact matrices [48]. Another driver of the disease dynamics is the mean generation time *T*_gen_, i.e., the average time between the exposures of an infector-infectee pair. It is a function of the mean latent period *T*_lat_ and the mean infectious period *T*_inf_. Since the period an individual spends in a certain compartment is typically not exponentially distributed [49, 50], we add realism to our model by further decomposing the infectious states in two substates, namely I_1_ and I_2_. The resulting infectious period is then gamma-distributed. Because of these two substates, the mean generation time *T*_gen_ in our model is [50]

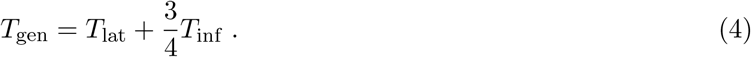

The detection process on the other hand will depend on the detectable period of the disease, *T*_det_, which corresponds to *T*_det_ = *T*_inf_ +*T*_post_, where *T*_post_ is the mean post-infectious period. Similarly to the infectious state, we decompose the post-infectious state in two substates, namely P_1_ and P_2_, but this does not affect the expression for *T*_det_. In Table 1, we provide the parameters’ ranges considered in this study and compare them with estimated ranges of SARS-CoV-2. Finally, another key characteristic of the disease dynamic is the doubling time *T*_2_. For the specific compartmental model considered here, it can obtained from the following implicit nonlinear equation [49] relating ℛ_0_ to the *growth rate λ*:

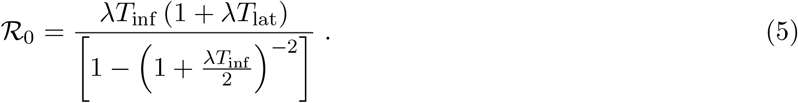

**Table 1:** Range of disease parameters explored in the main text and SI compared with estimates for the early transmission of SARS-CoV-2 (wild strain). ^†^Estimates combine mean generation time and serial interval.

| Parameter | Range considered | SARS-CoV-2 (wild strain) | References |
| --- | --- | --- | --- |
| Reproduction number | [1.5, 3] | [2, 3.5] | [43–46] |
| Mean generation time | [4, 36] days | [4, 7.5] days<sup>†</sup> | [44, 47] |
| Mean detectable period | [7.7, 22.7] days | [7, 22] days | [6, 29, 30] |

We solve this equation for *λ*, then we have *T*_2_ = ln 2*/λ*.

### Aircraft wastewater detection

In our model, a detectable individual passing through a sentinel site is detected with a probability *p*_det_ that depends on several factors, including the cadence and sampling of airport wastewater surveillance, the duration of the flight, the diverse sociodemographic profiles of the passengers [16], etc. In our analysis, we assume that on average, the detection probability *p*_det_ is uniform across all inbound international flights arriving at any given sentinel site. To provide a rationale for the spectrum of detection rates examined in this study, we break down the probability into the following components *p*_det_ = *p*_lav_ × *p*_shed_ × *p*_sample_ where *p*_lav_ represents the likelihood that an individual will utilize the lavatory and consequently deposit detectable genetic traces of the pathogen in the wastewater, *p*_shed_ denotes the probability that a detectable individual is actively shedding the pathogen at levels sufficient for detection in the wastewater, and *p*_sample_ refers to the proportion of flights that are subjected to sampling at the sentinel airport.

The proportion of adult passengers defecating on flights, critical for estimating *p*_lav_, is surveyed to be less than 13% on short-haul and less than 36% on long-haul flights [16]. Further, *p*_shed_, the probability of detectable pathogen shedding in fecal matter, ranges between 30% and 60% for SARS-CoV-2 [16]. Assuming that all international flights undergo sampling (*p*_sample_ = 100%), the resulting detection probability (*p*_det_) is calculated to be between 12% and 22%, corresponding to a detection rate of about 16% on long-haul flights. This estimate is possibly a large underestimation for viruses like SARS-CoV-2, as individuals can leave genetic material in the wastewater without defecating [51], such as by disposing of a used tissue or spitting in the toilet. Indeed, previous studies [21] have shown an 83.7% accuracy in detecting COVID-19 on repatriation flights using wastewater analysis. Translating this value to an individual’s marginal detection probability (*p*_det_) is complex, as the number of COVID-19 cases per flight varied significantly, averaging 4.62 cases. Accounting for false-positive wastewater results, assuming each case had an equal probability to be detected, and a flat prior on *p*_det_, we find a median marginal detection rate of 51% (90% CI, 28–72). However, this value could be inflated, notably due to the persistent nature of fecal RNA shedding compared to respiratory shedding [52]. Given the varying estimates discussed earlier, we explore detection rates up to 32%. However, acknowledging that not all international flights are long-haul and that only a fraction (e.g., 25%) of flights might be sampled, *p*_det_ could be as low as 4%. Our sensitivity analysis presents findings across this full spectrum of *p*_det_.

### Probability generating function analytics

The mechanistic GLEAM model employs large-scale stochastic simulations, which are computationally intensive. To streamline our analysis, for most of the results in this paper, we utilize probability generating functions (PGFs) to efficiently extract the required analytic information from the data and model. PGFs are a standard tool in mathematical epidemiology [53, 54], and have found many applications, including the quantitative analysis of the risk of introduction of diseases [55–58].

PGFs are useful to *count* elements. Here we are counting individuals based on certain properties: their age, their location, and their epidemiological state. We define *s*_*σ*_ as the number of individuals of *type σ*. For instance, *s*_*σ*_ could represent the current number of latent individuals in a given location and a certain age, or the cumulative number of individuals of a certain age that have been detected on a particular travel route. We use the vector ***s*** to encapsulate all these numbers.

To capture the full stochastic evolution of the system, we need to characterize the probability distribution *P* (***s***, *t*). We encode this distribution with a multivariate PGF

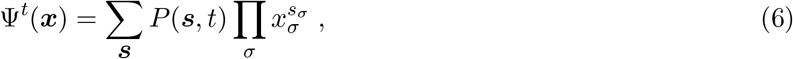

where the sum (product) runs over all possible values of ***s*** (*σ*) and each *x*_*σ*_ is a variable that acts as a placeholder—it does not mean anything and only serve to encode the probability distribution. The vector ***x*** encapsulates all these variables.

In the early stage, a structured metapopulation epidemic model like GLEAM can be described by a *multitype branching process* [59], in which case we can solve the PGF through the recursive equation

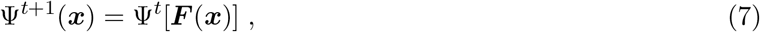

where ***F*** (***x***) is a vector of PGFs and each element *F*_*σ*_(***x***) is itself a PGF that characterizes the offspring distribution of an individual of type *σ*. For instance, the offspring distribution of an individual in the I_1_ state would give the probability that this individual generates a certain number of new latent individuals of each type through infections at the next time steps and the probability that this individual transitions to the I_2_ state. Computing the full distribution *P* (***s***, *t*) is generally out of reach—the number of terms explodes combinatorially. However, computing marginal or joint distributions for a few *observables*, like the total number of individuals in a particular state or the cumulative number of detections, is possible (see SI Sec. 1).

Altogether, the recursive evaluation of PGFs and their numerical inversion to recover probability distributions represents a very efficient computational alternative to Monte-Carlo simulations of GLEAM. Most notably, scanning different initial conditions is computationally cheap, since in Eq. (7), the PGF Ψ^0^(***x***) specifying the initial conditions is evaluated at the *end* of the recursion. This crucially allows us to extract distributions of observables, like the time to first detection, assuming the epidemic could have started from any of the 3200+ subpopulations of our model, a task that would be prohibitive with a purely simulation-based framework. See Sec. 1 of the SI for an in-depth description and characterization of the PGF methodology.

### WWSN optimization algorithms

The heuristic optimization of global WWSNs selects sentinel sites based on their rankings according to the following measures. Let *N*_*l→ν*_ be the number of individuals per day who will travel and arrive at airport *ν* on an international flight, either as a final destination or for a connection. The flows of international passengers generate a weighted bipartite network connecting international airports *ν* to subpopulations *l*. We can therefore rank airports based on their *volume* of international travel,

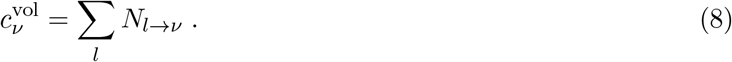

A second ranking measure is based on each airport traffic *entropy*, defined as:

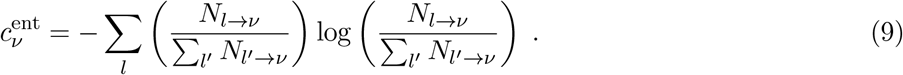

This expression is also known as Shannon’s diversity index. This measure favors airports with a broad and homogeneous coverage of the different subpopulations.

A more refined optimization algorithm aims at minimizing the mean time to first detection of an epidemic, averaged over all potential origins. We can assign an arbitrary prior probability *P* (*l*) for location *l* to be the origin of an epidemic, resulting in the following objective function

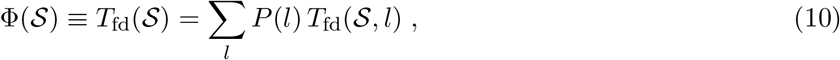

where *T*_fd_(𝒮, *l*) is the mean time to first detection, assuming the epidemic started in subpopulation *l* and that the WWSN consists of the set of sentinel airports 𝒮. This corresponds to the posterior mean of *T*_fd_ over all locations, which is proportional to *E*(𝒮), the excess time to first detection. For the global optimization, we use *P* (*l*) = const. ∀*l*, i.e., all subpopulations are equiprobable source. For the targeted optimization, we use *P* (*l*) = const. for the locations in the targeted region and *P* (*l*) = 0 otherwise.

We conjecture that −Φ(𝒮) is a *monotone submodular set function* [60]. We proved this statement in Sec. 2 of the SI for a very accurate approximation of −Φ(𝒮), but the exact case remains to be proven. Monotone submodular functions have desirable properties when it comes to discrete optimization problems. While minimizing Φ(𝒮) (equivalently maximizing −Φ(𝒮)) is an NP-hard problem, we have a guarantee on the performance of a *greedy* optimization algorithm—there exists an upper bound on the value of Φ(𝒮) obtained through this approach [60]. Most importantly, it is known that in practice, a greedy algorithm will find a solution that is very close to the optimal one. Consequently, to minimize the objection function Eq. (10), we use the following greedy optimization scheme:

1. define an initial set 𝒮 (can be empty);
2. for each airport *ν* ∉ 𝒮, compute Φ(𝒮 ∪ {*ν*});
3. update the set 𝒮 ← 𝒮 ∪ {*ν*^*⋆*^}, where *ν*^*⋆*^ is the sentinel airport that minimizes the objective function;
4. repeat steps 2-3 until a desired number of sentinels is reached.

The ‘greedy’ name comes from the fact that we are successively choosing a locally optimal choice at each stage of the algorithm (here in step 3).

## Data Availability

Proprietary airline data are commercially available from the Official Aviation Guide (https://www.oag.com/) and International Air Transport Association (https://www.iata.org/) databases. The mechanistic GLEAM model is publicly available at http://www.gleamviz.org/.

## Competing interests

SVS is a paid consultant at Verily. The authors declare no other relationships or activities that could appear to have influenced the submitted work.

## Acknowledgements

This work was in part supported by the Bill & Melinda Gates Foundation INV-058220. Under the grant conditions of the Foundation, a Creative Commons Attribution 4.0 Generic License has already been assigned to the Author Accepted Manuscript version that might arise from this submission. We acknowledge support from the CDC-75D301Cl4810 contract and the cooperative agreement CDC-RFA-FT-23-0069 from the CDC’s Center for Forecasting and Outbreak Analytics. GS acknowledges financial support from the Fonds de recherche du Québec – Nature et technologies (project 313475). LHD is supported by the National Institutes of Health 2P20GM125498-06 Centers of Biomedical Research Excellence Award. AA acknowledges support from the Natural Sciences and Engineering Research Council of Canada (projects 2019-05183 and 2024-05626). The findings and conclusions in this study are those of the authors and do not necessarily represent the official position of the funding agencies, the CDC, or the U.S. Department of Health and Human Services of the United States.

## Footnotes

1 These were the available air-travel networks at the beginning of the COVID-19 pandemic, and the one that was used to generate stochastic simulations from GLEAM at the time.

