## Supplementary Information for "Optimization and performance analytics of global aircraft-based wastewater surveillance networks"

Guillaume St-Onge[1,2], Jessica T. Davis[1], Laurent Hébert-Dufresne[3,4], Antoine Allard[3,4], Alessandra Urbinati[1], Samuel V. Scarpino[5,6,7], Matteo Chinazzi[1,2], and Alessandro Vespignani[1]

[1] *Laboratory for the Modeling of Biological and Socio-technical Systems, Northeastern University, Boston, MA 02115, USA*
[2] *The Roux Institute, Northeastern University, Portland, ME 04101, USA*
[3] *Vermont Complex Systems Center, University of Vermont, Burlington, VT 05401, USA*
[4] *Département de physique, de génie physique et d'optique, Université Laval, Québec City, QC G1V 0A6, Canada*
[5] *Institute for Experiential AI, Northeastern University, Boston, MA 02115, USA*
[6] *Network Science Institute, Northeastern University, Boston, MA 02115, USA*
[7] *Santa Fe Institute, Santa Fe, NM 87501, USA*

August 2, 2024

**4 Airport table for the sentinel surveillance system** **24**

#### 1 Model description

##### 1.1 Global Epidemic and Mobility Model

The GLEAM (Global Epidemic and Mobility) model is a stochastic epidemic metapopulation framework that incorporates age-based contact matrices and data on human mobility. The approach has been documented in previous publications [1, 2] and has been used to study the spread of diseases such as Ebola [3], Zika [4], and COVID-19 [5, 6]. The model uses a Voronoi tessellation to create a metapopulation network with over 3,200 subpopulations, covering areas of the globe inhabited by humans. These subpopulations are anchored around major transport hubs like airports and are themselves divided into cells measuring around 25 x 25 kilometers, equivalent to 15 x 15 arc minutes.

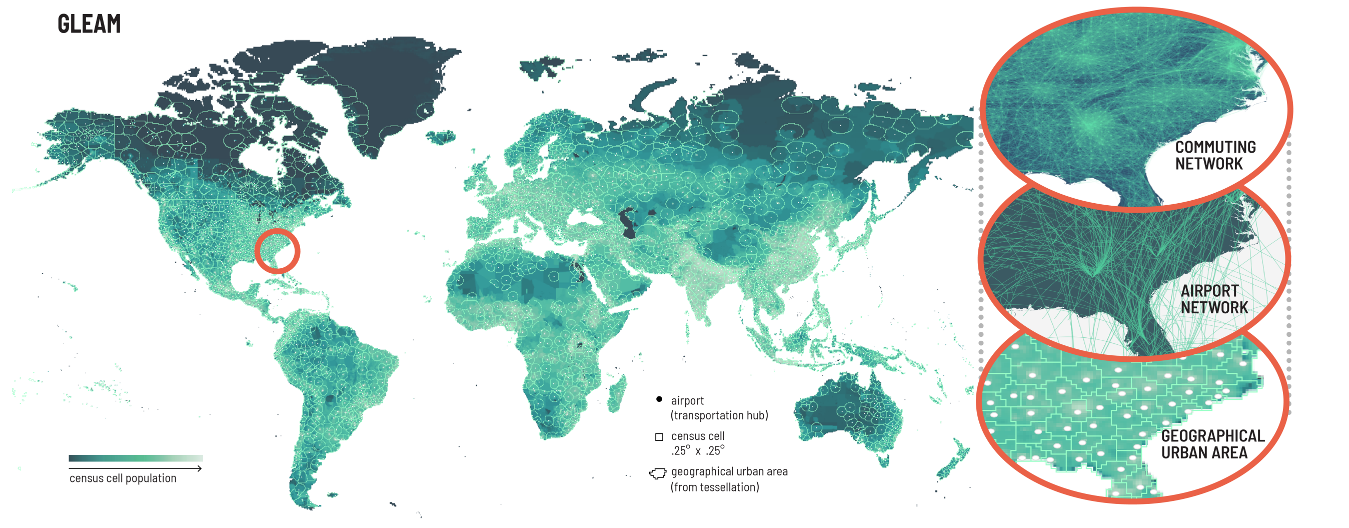

**Figure S1**: Schematic representation of GLEAM. (left) Voronoi tessellation of the globe centered around transportation hubs (airports) creating each of the 3200+ geographical units that we refer as subpopulations. Each subpopulation is constructed from census cells of approximately 25km × 25km. (right) For a particular region, we illustrate the subpopulations and the two mobility layers— air travel (long-range) and commuting (short-range).

In addition, the model integrates cell-level population data [7] and subpopulation-level age-specific contact patterns using the contact matrices developed in [8]. Here, we consider individuals divided into 5 age groups: [0-4, 5-17, 18-49, 50-64, 65+].

The individual subpopulations are connected through a human mobility layer that combines both short-range (i.e., commuting) and long-range (i.e., flights) mobility data. Commuting data is sourced from the Offices of Statistics for 30 countries on 5 continents. To harmonize the varying spatial resolutions of commuting data across different countries and to address data availability gaps, the short-range mobility layer is synthetically generated where necessary. This is achieved by relying on the "gravity law" [1, 9, 10], which is calibrated on the available data. Air-travel data from the Official Aviation Guide (OAG) and IATA databases is used to build an origin-destination network, incorporating connecting flight information. The network provides daily passenger flows between airports globally, which we systematically map and aggregate at the subpopulation level. Figure S1 displays the geographical resolution of the model for selected regions, illustrating both the short-range and long-range mobility networks and the global population structure.

#### Compartmental model

The synthetic world created by the human mobility layer couples the epidemic dynamics unfolding within each subpopulation. To model the infection process, we adopt an extended SLIR-like model in which individuals are either susceptible (S), latent (L), infectious (I), post-infectious (P), or removed (R). We further subdivide the infectious and post-infectious compartments into two stages, namely $I_1$, $I_2$, $P_1$, and $P_2$ to more realistically model the timing of the disease progression [11, 12]. Susceptible individuals become latent through interactions with infectious individuals, at a rate $\Lambda$, the *effective force of infection*, which depends on the age of the susceptible individual but also on the whole state of the system, and thus varies in time. Latent individuals progress to the first infectious stage at a constant rate inversely proportional to the mean latent period $\eta \equiv T_{\text{lat}}$. Infectious individuals in the first stage $I_1$ progress to the second stage $I_2$ at a rate $\mu \equiv 2 \times T_{\text{inf}}^{-1}$, where $T_{\text{inf}}$ is the mean infectious period; the process is identical for the

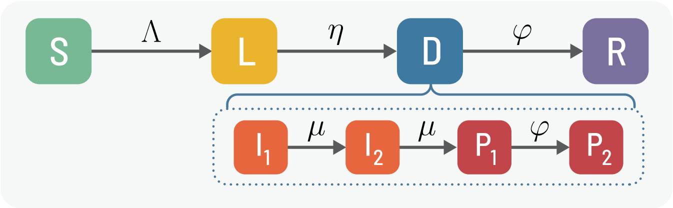

**Figure S2**: Compartmental model for wastewater surveillance at airports.

transition of $I_2$ individuals to $P_1$. Similarly, post-infectious individuals in the first stage ($P_1$) progress to the second stage $P_2$ at a rate $\varphi \equiv 2 \times T_{\text{post}}^{-1}$, where $T_{\text{post}}$ is the mean post-infectious period, then progress to the recovered stage at the same rate $\varphi$. The post-infectious period is the length of time that an individual can still shed the virus and remain detectable through wastewater, but not generate any more new infections. Once an individual is in the removed compartment, it can no longer be detected. Here we assume only infectious and post-infectious individuals can be detected through wastewater, which is why we regroup them in a *detectable* (D) meta-compartment. The various transitions and their rates are portrayed in Fig. S2.

Because of the subdivision of the infectious and post-infectious states, the infectious and post-infectious periods are gamma-distributed, while the latent period is exponentially distributed. The *generation time*, the time between the exposures of an infector-infectee pair, will also be gamma-distributed with mean $T_{\text{gen}}$ expressed as [11, 12]

$$T_{\text{gen}} = T_{\text{lat}} + \left( \frac{n_{\text{inf}} + 1}{2n_{\text{inf}}} \right) T_{\text{inf}} \ , \tag{1}$$

where $n_{\text{inf}}$ is the number of infectious states—in our model $n_{\text{inf}} = 2$. Similarly, the *detectable period*—the length of time an individual can be detected through wastewater—is gamma-distributed, with mean $T_{\text{det}} = T_{\text{inf}} + T_{\text{post}}$.

##### Stochastic simulation of the transmission and mobility dynamics

With the mobility data layers and the disease dynamics defined, the number of individuals in each compartment $c$, age bracket $a$, and subpopulation $l$ follows a discrete and stochastic dynamical equation that reads as

$$X_l^{[c,a]}(t + \Delta t) - X_l^{[c,a]}(t) = \Delta X_l^{[c,a]} + \Omega_l([c,a]) \tag{2}$$

where the term $\Delta X_l^{[c,a]}$ represents the changes induced by the disease dynamics and $\Omega_l([c,a])$ represents the variations due to air travel. In this study, each day is subdivided in $m = 12$ time steps, i.e., $\Delta t = 1/m$ days. While the disease dynamics part $\Delta X_l^{[c,a]}$ is applied at every of these time steps, the variations due to traveling $\Omega_l([c,a])$ are introduced daily. The latter $\Omega_l([c,a])$ is directly extracted from a multinomial distribution associated with the age-specific probability of travel per day, as defined by the global air-travel network.

The variation $\Delta X_l^{[c,a]}$ is determined by summing over all transitions in and out of the disease compartment $c$ for the age group $a$,

$$\Delta X_l^{[c,a]} = \sum_{[c',a]} \left\{ -\mathcal{D}_l([c,a],[c',a]) + \mathcal{D}_l([c',a],[c,a]) \right\} , \tag{3}$$

where $\mathcal{D}_l([c,a],[c',a])$ represents the number of transitions from $[c,a]$ to $[c',a]$ during the time interval $\Delta t$. For all spontaneous transitions, like from latent to infectious, $\mathcal{D}_l([c,a],[c',a])$ is simply extracted from a binomial distribution.

The generation of new infections is a more complex procedure. First, it hinges on the age-structured contacts matrix $\mathbf{C}$, which gives the expected number of contacts per day between each age pair $(a,a')$ in a given location $l$. We consider interactions in four social settings: contacts at school ($\mathbf{C}_{school}$), workplace ($\mathbf{C}_{work}$), home ($\mathbf{C}_{home}$), and in the general community ($\mathbf{C}_{community}$), which are linearly combined to create $\mathbf{C}$, as defined in Ref. [8]. Second, the mobility due to the commuting flows is also taken into account using a time scale separation approximation, as detailed in Ref. [1]. Altogether, these two factors contribute to the effective force of infections $\Lambda([l,a])$ acting on susceptible individuals in subpopulation $l$ and age $a$, resulting in new transitions of the form $\mathcal{D}_l([c,a],[c',a])$.

Initial conditions are established by defining the quantity and location of individuals who can spread the infection. Subsequently, GLEAM tracks the number of individuals in each disease compartment for every subpopulation over time. Please see Ref. [2] for a more in-depth discussion of the simulation framework.

###### Wastewater detection at airports

We assume that a subset of all airports $\mathcal{S} = \{\nu_1, \nu_2, \dots\}$—*sentinels*—monitor the wastewater of incoming *international* aircrafts. Consequently, each detectable international traveler passing through a sentinel has a probability $p_{\text{det}}$ of leading to a detection. As detailed in the Methods section of the main text, the probability $p_{\text{det}}$ is a complex quantity combining multiple factors, such as sampling frequency of aircrafts, length of the flight, demography, etc. We settled on performing an extensive sensitivity analysis spanning the range 4–32%.

In GLEAM's stochastic simulation process, detections at sentinels are aggregated during the post-processing of the simulation data, which keeps track of all mobility-induced changes $\Omega_l([c,a])$. Since GLEAM produces origin-destination travel patterns at the level of subpopulations $l$, we extract from the global air-travel network the probability of detection $p_{l,l'}$ for each travel $l \to l'$. Each travel $l \to l'$ is associated with a set of potential airport *paths* of the form $\mathcal{P} = \nu_1 \to \nu_2 \to \cdots \to \nu_k$. If the path $\mathcal{P}$ contains an international flight $\nu \to \nu'$, where $\nu' \in \mathcal{S}$, then detectable individuals taking this path will be detected with probability $p_{\text{det}}$. If a path $\mathcal{P}$ contains $\mathcal{N}(\mathcal{P})$ such international flight to any sentinel, the probability of detection is $1-(1-p_{\text{det}})^{\mathcal{N}(\mathcal{P})}$. We therefore calculate

$$p_{l,l'} = \sum_{\mathcal{P}} P(\mathcal{P}|l \to l') \left[1-(1-p_{\text{det}})^{\mathcal{N}(\mathcal{P})}\right] , \tag{4}$$

where $P(\mathcal{P}|l \to l')$ is the probability an individual will take the path $\mathcal{P}$, given the travel $l \to l'$.

##### 1.2 Probability generating function framework

To characterize the early phase of an epidemic, we can map the transmission tree to a branching process, from which an efficient *probability generating function* (PGF) methodology can be leveraged [13–17]. This is a standard approach in mathematical epidemiology and has been used, among others, to characterize the time evolution of contagion on heterogeneous networks [18–20], and to quantify the risk of introduction and outbreaks in metapopulation models [21–26]. Here we leverage this methodology by mapping to a multitype branching process the age-structured, stochastic, metapopulation dynamics of GLEAM, the disease progression, and the wastewater surveillance at airports (see Fig. S3).

PGFs are used to encode discrete probability distributions with functions. In the present case, we want to encode the full distribution for the state of the epidemic at all times $t$—the number of days since the start of the epidemic. The state of the epidemic includes the number of individuals in each stage of the disease, of each age, and in each subpopulation, but also other quantities we want to "measure" (evaluate an associated probability distribution), like the cumulative number of exported cases and the cumulative number of detections on each origin-destination travel ending at a sentinel.

To introduce our formalism, let us define the multi-index $\alpha = (l,a)$ characterizing the location $l$ and the age $a$ of an individual—we refer to $\alpha$ as the *category* of an individual or agent. We define $\ell_\alpha$ as the number of latent individuals of category $\alpha$. Since we divide the number of infectious and post-infectious states into two stages ($I_1$,$I_2$,$P_1$, and $P_2$) we identify them by the numbers $i_{\alpha,1}$, $i_{\alpha,2}$, and $j_{\alpha,1}$,$j_{\alpha,2}$ respectively. To track the number of exported cases, we introduce $e_\alpha$ as the cumulative number of $\alpha$-agents, either latent or infectious, who were infected previously in another location—they were previously of another category—then traveled, thereby becoming of category $\alpha$. Finally, to track wastewater detections of detectable individuals (infectious or post-infectious) at sentinel airports, we define

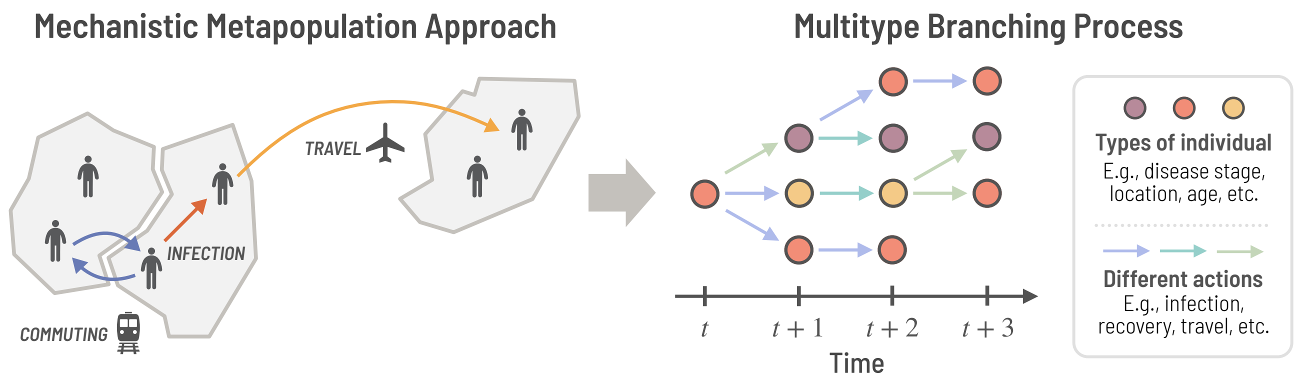

**Figure S3**: Mapping GLEAM to a multitype branching process. Infections happen within subpopulations—and across adjacent subpopulations when accounting for commuting—and agents can travel between the subpopulations. Individuals are distinguished by their *type*, which encapsulates everything that distinguishes them (like disease stage, age, and location). The early phase of an epidemic naturally takes the form of an event tree encoding all actions happening at each time step, including travel and transmission. This event tree is mathematically described via a multitype branching process.

$d_{\alpha,\alpha'}$ the cumulative number of detections for international travel $l \to l'$ resulting in a change of category $\alpha \to \alpha'$ for a given age group $a$. Note that we do not track susceptible or recovered individuals in this framework, since they do not play a significant role in the early phase of an outbreak. The whole state of the system is then described by the tuple of vectors $(\boldsymbol{\ell}, \boldsymbol{i}_1, \boldsymbol{i}_2, \boldsymbol{j}_1, \boldsymbol{j}_2, \boldsymbol{e}, \boldsymbol{d})$, where for instance $\boldsymbol{\ell} = [l_{\alpha_1}, l_{\alpha_2}, \dots]$.

To simplify the notation, we define the *state vector*

$$\boldsymbol{s} = [l_{\alpha_1}, l_{\alpha_2}, \dots, i_{\alpha_1,1}, i_{\alpha_2,2}, \dots, d_{\alpha_1,\alpha_2}, d_{\alpha_1,\alpha_3}, \dots] \equiv [s_1, s_2, \dots] \,, \tag{5}$$

as the concatenation of previous vectors, encoding all information about the system. Each component $s_\sigma$ of the vector identifies a number we keep track of (e.g., number of infectious of a specific category $\alpha$) and is associated with nodes of a specific color in Fig. S3; we refer to $\sigma$ as the *type* of each node in the multitype branching process representation of the epidemic and mobility dynamics.

Since the state vector is a random variable $\boldsymbol{s}_t$ at each time $t$, we write the probability of observing a particular state at time $t$ as $P(\boldsymbol{s}_t = \boldsymbol{s}) \equiv P(\boldsymbol{s}, t)$. Finally, we encode this distribution with a PGF (in the compact format) as

$$\Psi^t(\boldsymbol{x}) \equiv \sum_{\boldsymbol{s}} P(\boldsymbol{s}, t) \prod_{\sigma} x_\sigma^{s_\sigma} \,, \tag{6}$$

where the sum runs over all potential values of the vector $\boldsymbol{s}$ and each $x_\sigma$ is a dummy variable used to track each quantity $s_\sigma$.

If we unravel the vector $\boldsymbol{s}$, the PGF is written as

$$\Psi^t(\boldsymbol{z}, \boldsymbol{y}_1, \boldsymbol{y}_2, \boldsymbol{w}_1, \boldsymbol{w}_2, \boldsymbol{v}, \boldsymbol{u}) = \sum_{\boldsymbol{\ell}} \sum_{\boldsymbol{i}_1} \sum_{\boldsymbol{i}_2} \sum_{\boldsymbol{j}_1} \sum_{\boldsymbol{j}_2} \sum_{\boldsymbol{e}} \sum_{\boldsymbol{d}} P(\boldsymbol{\ell}, \boldsymbol{i}_1, \boldsymbol{i}_2, \boldsymbol{j}_1, \boldsymbol{j}_2, \boldsymbol{e}, \boldsymbol{d}, t) \prod_{\alpha,\alpha'} z_\alpha^{\ell_\alpha} y_{\alpha,1}^{i_{\alpha,1}} y_{\alpha,2}^{i_{\alpha,2}} w_{\alpha,1}^{j_{\alpha,1}} w_{\alpha,2}^{j_{\alpha,2}} v_\alpha^{e_\alpha} u_{\alpha,\alpha'}^{d_{\alpha,\alpha'}} \,, \tag{7}$$

where the vector of variables $(\boldsymbol{z}, \boldsymbol{y}_1, \boldsymbol{y}_2, \boldsymbol{w}_1, \boldsymbol{w}_2, \boldsymbol{v}, \boldsymbol{u})$ tracks the quantities $(\boldsymbol{\ell}, \boldsymbol{i}_1, \boldsymbol{i}_2, \boldsymbol{j}_1, \boldsymbol{j}_2, \boldsymbol{e}, \boldsymbol{d})$. For obvious reasons, we will favor the compact representation as much as possible, and use the unraveled vectors only when necessary to specify operations on a subset of the variables.

For a general multitype branching process, the solution is obtained by recursion [18]

$$\Psi^{t+1}(\boldsymbol{x}) = \Psi^t\left[\boldsymbol{F}(\boldsymbol{x})\right] \ , \tag{8}$$

where $F(\boldsymbol{x}) = [F_1(\boldsymbol{x}), F_2(\boldsymbol{x}), \dots]$ is a vector of PGFs, and each PGF $F_\sigma(\boldsymbol{x})$ characterizes the *offspring* distribution at the next time step for each node of type $\sigma$. For instance, assuming $\sigma$ identifies infectious $\alpha$-agents in the first stage ($I_1$), each individual could lead to a certain number of new latent individuals (through transmission), an $\alpha$-agent in the second stage ($I_2$) through disease progression, detection at airports through travel, and so forth. Each possible combination of offspring is encoded in a multivariate PGF, $F_\sigma(\boldsymbol{x})$, similar to Eq. (6).

We now decompose the vector of offspring PGFs $\boldsymbol{F}(\boldsymbol{x})$ by describing separately the reaction phase—modeling disease transmission and progression—, and the mobility phase of GLEAM in terms of vectors of PGFs $\boldsymbol{R}(\boldsymbol{x})$ and $\boldsymbol{M}(\boldsymbol{x})$ respectively. From a similar argument justifying Eq. (8), the vector of offspring PGFs is the composition of the PGFs for each phase [18], i.e., $\boldsymbol{F}(\boldsymbol{x}) = \boldsymbol{M}(\boldsymbol{R}(\boldsymbol{x}))$. The order of the composition indicates that air travel is happening *before* the reaction phase, in line with the convention in GLEAM.

##### Reaction phase: disease transmission and progression

While the PGF framework Eq. (8) is defined at a daily resolution, we model the reaction phase at a finer temporal resolution by dividing each day into $m = 12$ time periods of duration $\Delta t = 1/m$, as in the GLEAM simulation procedure. This provides a more realistic description of the intraday dynamics, especially for rapidly evolving epidemics, but one could work at any temporal resolution without significantly affecting the results. Consequently, the vector of PGFs $\boldsymbol{R}(\boldsymbol{x})$ for the reaction phase itself corresponds to the $m$-th composition of the vector of PGFs $\boldsymbol{r}(\boldsymbol{x})$, i.e.,

$$\boldsymbol{R}(\boldsymbol{x}) = \underbrace{\boldsymbol{r}(\boldsymbol{r}(\dots\boldsymbol{r}(}_{m\text{ times}}\boldsymbol{x})\dots)) \ . \tag{9}$$

Each transition in the disease progression has a rate: latent individuals become infectious at rate $\eta$, infectious individuals in the first stage transition to the second stage (and then in the post-infectious stage) at rate $\mu$, post-infectious individuals in the first stage transition to the second stage (and then in the removed stage) at rate $\varphi$. Also, infectious $\alpha$-agents (at any stage) interact and transmit the disease to $\alpha'$-susceptible individuals at rate $\beta_{\alpha,\alpha'}$, which is constructed from the age-structured contact matrix $\mathbf{C}$ and take into account commuting.

Considering all these possible transitions, we report in Table S1 the offspring PGF associated to a single step of the reaction phase, $r_\sigma(\boldsymbol{x})$, for each type of node. Disease progression transitions are represented by multinomial PGFs, while disease transmission (for infectious individuals only) is represented by a multivariate Poisson PGF. The multivariate Poisson PGF corresponds to an infinite-size subpopulation approximation for binomial draws used in GLEAM simulations for the infection process. Note that the offspring PGFs for elements of type $\sigma$ identifying a cumulative number of exported individuals or a cumulative number of wastewater detections at airports are simply the identity PGF, i.e., $f(x) = x$. Indeed, these quantities do not generate new infections or transition to other states, they only serve to keep track of a sum [18].

##### Mobility phase: air-travel and detection at airports

Each day, latent, infectious, and post-infectious $\alpha$-agents in location $l$ move to a new subpopulation $l'$ of multi-index $\alpha'$ with probability $m_{\alpha,\alpha'}$ and stay with probability $m_{\alpha,\alpha}$; agents that move and are detectable (infectious or post-infectious) are detected with probability $p_{\alpha,\alpha'} = p_{l,l'}$, calculated in Eq. (4). Similarly to the reaction phase, we report in Table S2 the offspring PGF associated with the mobility phase, $M_\sigma(\boldsymbol{x})$, for each type of node. Except for the types identifying exported individuals and wastewater detections, $M_\sigma(\boldsymbol{x})$ is a multinomial PGF.

**Table S1**: Offspring PGFs associated with a single step of the reaction phase for each element of the state vector. The parameters $\eta, \mu, \phi$ correspond to spontaneous transition rates between the disease stages (see Fig. S2). The parameter $\beta_{\alpha,\alpha'}$ is the rate at which infectious $\alpha$-agents interact and transmit the disease to $\alpha'$-susceptible individuals.

| Element identified by the type $\sigma$ | Reaction phase PGF $r_\sigma(\boldsymbol{x})$ |
|---|---|
| Latent individuals of category $\alpha$ | $y_{\alpha,1}\eta\Delta t + z_\alpha(1-\eta\Delta t)$ |
| Infectious individuals of category $\alpha$ in the first stage | $\exp\left[\sum_{\alpha'}\beta_{\alpha,\alpha'}(z_{\alpha'}-1)\Delta t\right]\left[y_{\alpha,2}\mu\Delta t + y_{\alpha,1}(1-\mu\Delta t)\right]$ |
| Infectious individuals of category $\alpha$ in the second stage | $\exp\left[\sum_{\alpha'}\beta_{\alpha,\alpha'}(z_{\alpha'}-1)\Delta t\right]\left[w_{\alpha,1}\mu\Delta t + y_{\alpha,2}(1-\mu\Delta t)\right]$ |
| Post-infectious individuals of category $\alpha$ in the first stage | $w_{\alpha,2}\varphi\Delta t + w_{\alpha,1}(1-\varphi\Delta t)$ |
| Post-infectious individuals of category $\alpha$ in the second stage | $\varphi\Delta t + w_{\alpha,2}(1-\varphi\Delta t)$ |
| Exported individuals of category $\alpha$ | $v_\alpha$ |
| Wastewater detections related to a change of category $\alpha\to\alpha'$ | $u_{\alpha,\alpha'}$ |

**Table S2**: Offspring PGFs associated with the mobility phase for each element of the state vector. The parameter $m_{\alpha,\alpha'}$ is the probability per day for $\alpha$-agents to move and become of category $\alpha'$; the parameter $p_{\alpha,\alpha'}$ is the probability of detection for detectable individuals if they move according to $\alpha\to\alpha'$.

| Element identified by the type $\sigma$ | Mobility phase PGF $M_\sigma(\boldsymbol{x})$ |
|---|---|
| Latent individuals of category $\alpha$ | $\sum_{\alpha'\neq\alpha} m_{\alpha,\alpha'}z_{\alpha'}v_{\alpha'} + m_{\alpha,\alpha}z_\alpha$ |
| Infectious individuals of category $\alpha$ in the first stage | $\sum_{\alpha'\neq\alpha} m_{\alpha,\alpha'}y_{\alpha',1}v_{\alpha'}(1-p_{\alpha,\alpha'}+p_{\alpha,\alpha'}u_{\alpha,\alpha'}) + m_{\alpha,\alpha}y_{\alpha,1}$ |
| Infectious individuals of category $\alpha$ in the second stage | $\sum_{\alpha'\neq\alpha} m_{\alpha,\alpha'}y_{\alpha',2}v_{\alpha'}(1-p_{\alpha,\alpha'}+p_{\alpha,\alpha'}u_{\alpha,\alpha'}) + m_{\alpha,\alpha}y_{\alpha,2}$ |
| Post-infectious individuals of category $\alpha$ in the first stage | $\sum_{\alpha'\neq\alpha} m_{\alpha,\alpha'}w_{\alpha',1}(1-p_{\alpha,\alpha'}+p_{\alpha,\alpha'}u_{\alpha,\alpha'}) + m_{\alpha,\alpha}w_{\alpha,1}$ |
| Post-infectious individuals of category $\alpha$ in the second stage | $\sum_{\alpha'\neq\alpha} m_{\alpha,\alpha'}w_{\alpha',2}(1-p_{\alpha,\alpha'}+p_{\alpha,\alpha'}u_{\alpha,\alpha'}) + m_{\alpha,\alpha}w_{\alpha,2}$ |
| Exported individuals of category $\alpha$ | $v_\alpha$ |
| Wastewater detections related to a change of category $\alpha\to\alpha'$ | $u_{\alpha,\alpha'}$ |

##### Initial conditions

The solution of Eq. (8) takes the form

$$\Psi^t(\boldsymbol{x}) = \Psi^0(\underbrace{\boldsymbol{F}(\boldsymbol{F}(\cdots\boldsymbol{F}(}_{t\text{ times}}\boldsymbol{x})\cdots))) \,. \tag{10}$$

The PGF $\Psi^0(\boldsymbol{x})$ specifies the initial condition. In all cases in this study, we specify a number $\ell$ and $i_1$ of initial latent and infectious ($\mathrm{I}_1$) individuals respectively. Their age is randomly selected according to the age distribution of the origin subpopulation $l$, which we represent by the conditional category distribution $P(\alpha|l)$. The PGF for the initial conditions then takes the form

$$\Psi^0(\boldsymbol{x}) = \left(\sum_\alpha P(\alpha|l)z_\alpha\right)^\ell \left(\sum_\alpha P(\alpha|l)y_{\alpha,1}\right)^{i_1} \,, \tag{11}$$

It is worth stressing that since $\Psi^0$ is applied *last* when evaluating Eq. (10), it is computationally inexpensive to test various initial conditions, which we use in this study to vary the origin of the outbreak.

##### Evaluation of probability distributions and cumulants

For a large number of multi-indices $\sigma$, evaluating the full joint distribution $P(\boldsymbol{s},t)$ is computationally prohibitive. However, it is not our goal: In general, we want to evaluate a marginal or a joint distribution of some *observables*, like $d_t$, the total number of detections at any sentinels by time $t$, which can be estimated from the PGF $\Psi^t(\boldsymbol{x})$. For instance, the PGF for the distribution $P(d_t = d)$, is

$$\zeta^t(x) \equiv \sum_d P(d_t = d) x^d = \Psi^t \left[\boldsymbol{A}(x)\right] \ , \tag{12}$$

where $\boldsymbol{A}(x)$ is a vector where $A_\sigma(x) = x$ if $\sigma$ identifies any of the cumulative detection at a sentinel for a specific pair of category, $d_{\alpha,\alpha'}$, and $A_\sigma(x) = 1$ otherwise. This allows us to implicitly sum over all possible combinations of $d_{\alpha,\alpha'}$ resulting in a total number of detection $d_t$. PGFs $\zeta^t(x,y)$ for joint distributions of observables are constructed in similar fashion.

For any observable $n_t$, we can extract the distribution $P(n_t)$ from its PGF $\zeta^t(x) = \sum_n P(n_t = n) x^n$ using the following identity

$$\begin{aligned} P(n_t = n) &= \frac{1}{n!} \left. \frac{\mathrm{d}^n}{\mathrm{d}x^n} \zeta^t(x) \right|_{x=0} \ , \\ &= \frac{1}{2\pi\rho^n} \int_0^{2\pi} \zeta^t\left(\rho e^{i\omega}\right) e^{-i\omega n} \mathrm{d}\omega \ , \end{aligned} \tag{13}$$

where $0 < \rho < 1$ is a free control parameter. In practice, we use the following discrete Fourier transform approximation (efficiently calculated using Fast Fourier Transform algorithms)

$$P(n_t = n) \approx \frac{1}{k\rho^n} \sum_{j=0}^{k-1} \zeta^t\left(e^{2\pi i j/k}\right) e^{-2\pi i j n/k} \ . \tag{14}$$

Note that it is possible to bound and control the error committed by choosing $k$ and a suitable $\rho$ value [27], making this approximation exact for all practical purposes. An analogous multidimensional discrete Fourier transform numerical solution is used for distributions of joint observables.

Finally, in some cases, it is more convenient and computationally efficient to extract a few cumulants—mean, variance, etc.—instead of the full distribution. Fortunately, the *cumulant generating function* (CGF) $K^t(x)$ for an observable is directly related to its PGF $\zeta^t(x)$ by the relation $K^t(x) = \ln \zeta^t(e^x)$ [28]. Cumulants are extracted from the CGF using a discrete Fourier transform procedure similar to Eq. (14), which is also directly generalizable to higher dimensions (joint cumulants).

**Minimal example: SIR dynamics in two subpopulations**

To illustrate the full PGF methodolology from end to end, let us consider a simpler, minimal, example of two subpopulations and a single age group. Therefore, we only have two categories $\alpha \in \{1,2\}$ associated to each subpopulation. We also consider a simpler compartmental model: individuals are either susceptible, infectious, or recovered; since in our framework we do not track susceptible and recovered individuals, we only need to track $i_1$ and $i_2$, the number of infectious individuals in each subpopulations. The probability distribution for the state of the epidemic at time $t$, $P(i_1, i_2, t)$, is encoded with the PGF

$$\Psi^t(\boldsymbol{x}) = \Psi^t(y_1, y_2) = \sum_{i_1=0}^{\infty} \sum_{i_2=0}^{\infty} P(i_1, i_2, t) y_1^{i_1} y_2^{i_2} \ , \tag{15}$$

where $y_1$ and $y_2$ are placeholder variables tracking $i_1$ and $i_2$ respectively.

We assume that the epidemic starts at time $t = 0$ with 1 infectious individuals in subpopulation 1. From Eq. 11, this implies $\Psi^0(y_1, y_2) = y_1$. Let us now consider the evolution of the system for a single time step. To simplify the expressions, we use $m = 1$ ($\Delta t = 1$) for the temporal scale of the reaction phase. The offspring PGFs associated with the mobility phase are written as follows

$$\boldsymbol{M}(y_1, y_2) = \begin{bmatrix} M_1(y_1, y_2) \\ M_2(y_1, y_2) \end{bmatrix} = \begin{bmatrix} m_{1,1} y_1 + m_{1,2} y_2 \\ m_{2,1} y_2 + m_{2,2} y_2 \end{bmatrix} = \begin{bmatrix} m_{1,1} & m_{1,2} \\ m_{2,1} & m_{2,2} \end{bmatrix} \begin{bmatrix} y_1 \\ y_2 \end{bmatrix} \ . \tag{16}$$

We interpret $M_1(y_1, y_2) = m_{1,1}y_1 + m_{1,2}y_2$ as follows: an infectious individuals in subpopulation 1 has a probability $m_{1,1}$ to stay in subpopulation 1 and a probablity $m_{1,2} = 1 - m_{1,1}$ to move to subpopulation 2. A similar reasoning holds for $M_2(y_1, y_2)$.

The offspring PGFs associated with the reaction phase are written as follows

$$\boldsymbol{R}(y_1, y_2) = \begin{bmatrix} R_1(y_1, y_2) \\ R_2(y_1, y_2) \end{bmatrix} = \begin{bmatrix} \exp\{\beta_{1,1}(y_1 - 1)\} \exp\{\beta_{1,2}(y_2 - 1)\}\{\mu + (1 - \mu)y_1\} \\ \exp\{\beta_{2,1}(y_1 - 1)\} \exp\{\beta_{2,2}(y_2 - 1)\}\{\mu + (1 - \mu)y_2\} \end{bmatrix} . \tag{17}$$

We interpret $R_1(y_1, y_2)$ as follows: 1) An infectious individual in subpopulation 1 will generate a number of secondary infections in subpopulation 1, distributed according to a Poisson distribution with mean $\beta_{1,1}$, to which corresponds the PGF $\exp\{\beta_{1,1}(y_1 - 1)\}$. 2) If we take into account commuting, i.e., $\beta_{1,2} > 0$, an infectious individual in subpopulation 1 will also generate secondary infections in subpopulation 2, with PGF $\exp\{\beta_{1,2}(y_2 - 1)\}$. 3) An infectious individual in subpopulation 1 will either recover with probablity $\mu$ or remain infectious with probability $1 - \mu$, resulting in the PGF $\mu + (1 - \mu)y_1$. Since these 3 kind of events are *independent*, the PGF for the sum is the product of the individual PGFs [17], resulting in the expression for $R_1(y_1, y_2)$. A similar reasoning holds for $R_2(y_1, y_2)$.

Combining the mobility and reaction phases, the offspring PGFs $\boldsymbol{F}(\boldsymbol{x}) = \boldsymbol{M}(\boldsymbol{R}(\boldsymbol{x}))$ for infectious individuals in both subpopulations are

$$\boldsymbol{F}(y_1, y_2) = \begin{bmatrix} F_1(y_1, y_2) \\ F_2(y_1, y_2) \end{bmatrix} = \begin{bmatrix} M_1(R_1(y_1, y_2), R_2(y_1, y_2)) \\ M_2(R_1(y_1, y_2), R_2(y_1, y_2)) \end{bmatrix} = \begin{bmatrix} m_{1,1} & m_{1,2} \\ m_{2,1} & m_{2,2} \end{bmatrix} \begin{bmatrix} R_1(y_1, y_2) \\ R_2(y_1, y_2) \end{bmatrix} . \tag{18}$$

Using the recursion in Eq. (8) and the offspring PGFs above, we are able to calculate the PGF for the state of the epidemic at time $t = 1$

$$\Psi^1(y_1, y_2) = \Psi^0(F_1(y_1, y_2), F_2(y_1, y_2)) = F_1(y_1, y_2) = m_{1,1}R_1(y_1, y_2) + m_{1,2}R_2(y_1, y_2) . \tag{19}$$

We interpret Eq. (19) as follows: with probability $m_{1,1}$, the index case will stay in the first subpopulation, and generate secondary infections as prescribed by $R_1(y_1, y_2)$, or travel to subpopulation 2 with probability $m_{1,2}$ and generate secondary infections as prescribed by $R_2(y_1, y_2)$.

From $\Psi^1(y_1, y_2)$, we can estimate any distributions of *observables* for the state of the epidemic at time $t = 1$. For instance, if we compose $\Psi^1$ with $\boldsymbol{A}(x) = [A_1(x), A_2(x)]^\top = [x, x]^\top$ to get the PGF

$$\zeta^1(x) = \Psi^1(x, x) = m_{1,1}R_1(x, x) + m_{1,2}R_2(x, x) , \tag{20}$$

then $\zeta^1(x)$ encodes the distribution for the total number of infectious in both subpopulations, which can be recovered numerically through Eq. (14).

Altogether, this minimal example with the SIR model in two subpopulations illustrates the full PGF methodology: We start from a PGF describing the initial conditions, $\Psi^0(\boldsymbol{x})$, we define the vector of offspring PGFs $\boldsymbol{F}(\boldsymbol{x})$, using Eq. (8) we get the PGF for the state of the epidemic at a later time $\Psi^t(\boldsymbol{x})$, and finally we use it to get a PGF for the disribution of an observable $\zeta^t(x)$, like the total number of infectious individuals. For the compartmental model in Fig. S2 and the metapopulation network of GLEAM, the dimension of the PGFs and equations is much larger, but the procedure is exactly the same.

#### 2 Time to first detection for a global wastewater surveillance system at airports

An important quantity introduced in the main text is the time to first detection, $t_{\rm fd}$, with distribution

$$P(t_{\rm fd} = t) = P(d_{t-1} < 1, d_t \geq 1) . \tag{21}$$

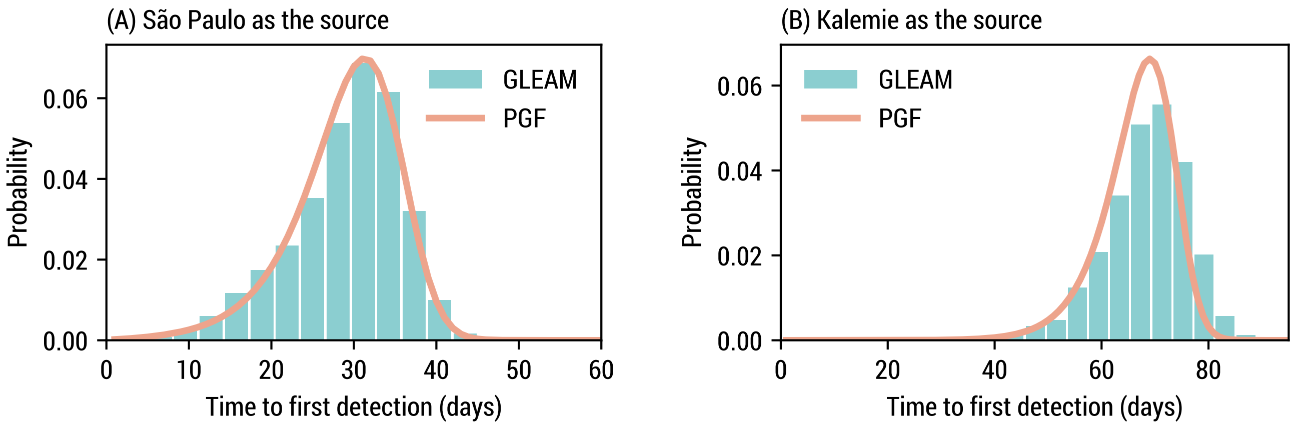

**Figure S4**: Comparison of the time to first detection. We use the same model, parametrization, and baseline WWSN as in Fig. 1. (A) The epidemic starts in São Paulo. GLEAM estimates are based on 1135 simulation runs. The mean time to first detection is 29.98 (SE, 0.19) days for GLEAM and 29.97 days using the PGFs. (B) The epidemic starts in Kalemie. GLEAM estimates are based on 1590 simulation runs. The mean time to first detection is 68.47 (SE, 0.20) days for GLEAM and 66.46 days using the PGFs.

We can simplify the joint distribution on the right-hand side. Note that the statements $A \equiv d_{t-1} < 1$ and $B \equiv d_t \geq 1$ are Boolean random variables. We can write the probability $P(A, B) = P(A) - P(A, \neg B)$. Since $P(\neg A, \neg B) = 0$ because it is impossible, then $P(A, \neg B) = P(\neg B)$. Therefore,

$$P(d_{t-1} < 1, d_t \geq 1) \equiv P(A) - P(\neg B) = P(d_{t-1} < 1) - P(d_t < 1) = P(d_{t-1} = 0) - P(d_t = 0) \ , \tag{22}$$

which is straightforward to evaluate using the PGFs. Let us emphasize that the time to first detection is independent of the wastewater methodology being used (pooled sampling or individual aircraft sampling), in the sense that it does not matter if more than one detectable individual contributed to a positive test at a sentinel. It is also worth highlighting the close connection with the extensive line of work on the *arrival time* of a disease in metapopulation networks [29–33].

To complement Fig. 1 in the main text, we validate in Fig. S4 the PGF solutions with GLEAM simulations for the full distribution of $t_{\text{fd}}$ for two epidemic origins. In Fig. S4(A), the PGF prediction accurately reproduces the distribution, with $T_{\text{fd}}$, the mean time to first detection, being within less than 1 standard error (SE) of the simulation estimate. In Fig. S4(B), there are approximately 2 days of difference between the two estimates of $T_{\text{fd}}$ (i.e., 3% difference), which is due to finite subpopulation effects not taken into account by the PGFs. Note that Kalemie is a subpopulation for which it takes a long time to detect with this surveillance system and therefore accentuates the discrepancy.

In Fig. 2 of the main text, we present results for $T_{\text{fd}}$ aggregated over continents; in Fig. S5, we present a similar same analysis, but aggregating over statistical subregions in Africa and Asia. We again observe broad heterogeneity of the mean time to first detection at this geographical scale in all subregions.

An important surveillance network introduced in the main text is the complete WWSN, where all airports act as surveillance sites. In Fig S6, we show $T_{\text{fd}}$ from every potential origin in the world for the complete WWSN. We note the similarity with the global map obtained in Figs. 1. The mean time to first detection is still very heterogeneous, with some locations taking a week and others as much as 100 days before a first detection by the WWSN. To better explain the source of this heterogeneity, in Fig. S7, we show the relation between the probability per day of traveling to an international destination for an individual in each location and $T_{\text{fd}}$. There is a clear negative correlation between

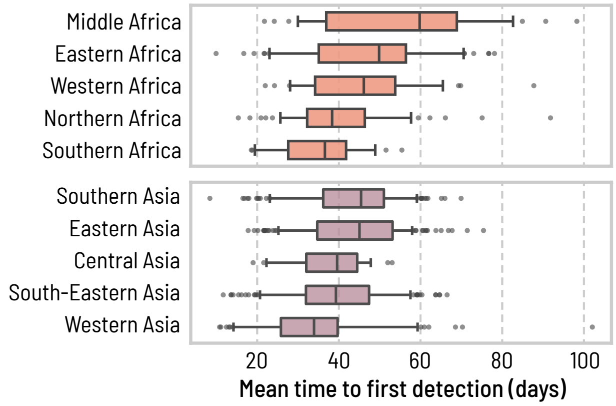

| subregion | min | P5 | median | mean | P95 | max |
|---|---|---|---|---|---|---|
| M. Africa | 21.8 | 28.2 | 59.8 | 55.8 | 84.5 | 98.3 |
| E. Africa | 9.9 | 23.0 | 49.9 | 47.2 | 70.5 | 78.1 |
| W. Africa | 22.0 | 28.0 | 46.1 | 46.2 | 67.2 | 87.7 |
| N. Africa | 15.4 | 24.5 | 38.4 | 40.1 | 58.7 | 91.8 |
| S. Africa | 18.6 | 19.1 | 36.6 | 35.6 | 50.2 | 55.4 |
| S. Asia | 8.4 | 23.1 | 45.4 | 43.6 | 59.1 | 69.9 |
| E. Asia | 17.8 | 24.9 | 45.0 | 43.9 | 58.1 | 75.4 |
| C. Asia | 19.0 | 22.1 | 39.6 | 37.7 | 48.6 | 53.0 |
| S.-E. Asia | 11.7 | 20.6 | 39.3 | 39.6 | 57.7 | 66.5 |
| W. Asia | 10.8 | 14.2 | 33.9 | 34.1 | 59.3 | 102.1 |

**Figure S5**: Heterogeneity of the time to first detection within statistical subregions. We aggregate the mean time to first detection $T_{\text{fd}}$ obtained in Fig. 1 over statistical subregions as defined by the United Nations geoscheme in Africa (Middle, $n = 45$; Eastern $n = 121$; Western $n = 52$; Northern $n = 89$; Southern $n = 31$) and Asia (Southern $n = 205$; Eastern $n = 269$; Central $n = 37$; South-Eastern $n = 236$; Western $n = 120$). The center line of the box plot indicates the median, the box covers the interquartile range, the whiskers cover the 90% central prediction interval (P5–P95), and black dots correspond to outliers outside this interval. Numerical values for some of the statistics of the mean time to first detection are reported in the table on the right.

the two—a higher probability lead to a faster detection—, but other factors are important. For instance, for some subpopulations, international travel is not directly possible—an individual must first travel to another subpopulation within the same country before moving abroad. In other words, one must take into account additional seeding events through importations or commuting to fully characterize the time to first detection.

#### 2.1 Variation with the disease natural history

Changing the disease's natural history affects significantly $T_{\text{fd}}$, as can be seen in Fig. 3 in the main text. For the sake of completeness, we also show in Fig. S8 a similar analysis, where instead of varying generation time via the latent period, we do it via the infectious period, keeping the detectable period fixed. We obtain similar results for the mean time to first detection as in Fig. 3.

By changing the generation time or the reproduction number, we ultimately change the growth rate $\lambda$ of the epidemic—or equivalently the doubling time $T_2 \equiv \ln 2/\lambda$. For the model we consider, the growth rate can be obtained by solving the following implicit nonlinear equation [11]:

$$\mathcal{R}_0 = \lambda T_{\text{inf}} \frac{\left(1 + \frac{\lambda T_{\text{lat}}}{n_{\text{lat}}}\right)^{n_{\text{lat}}}}{\left[1 - \left(1 + \frac{\lambda T_{\text{inf}}}{n_{\text{inf}}}\right)^{-n_{\text{inf}}}\right]} \,, \tag{23}$$

where $n_{\text{lat}}$ and $n_{\text{inf}}$ are the numbers of latent and infectious states respectively (here we use $n_{\text{lat}} = 1$ and $n_{\text{inf}} = 2$).

In Fig. 3 of the main text, we show that for all practical purposes, the following relationship holds:

$$\frac{T_{\text{fd}}}{T_2} + \log_2 T_2 \approx \text{const.} \tag{24}$$

Another way to interpret this equation is that changing the doubling time from $T_2$ to $\hat{T}_2$ ultimately amounts to a linear transformation of the form

$$\hat{T}_{\text{fd}} = aT_{\text{fd}} + b \quad ; \quad a = \left(\frac{\hat{T}_2}{T_2}\right) \quad ; \quad b = \hat{T}_2 \log_2 \left(\frac{T_2}{\hat{T}_2}\right) \,, \tag{25}$$

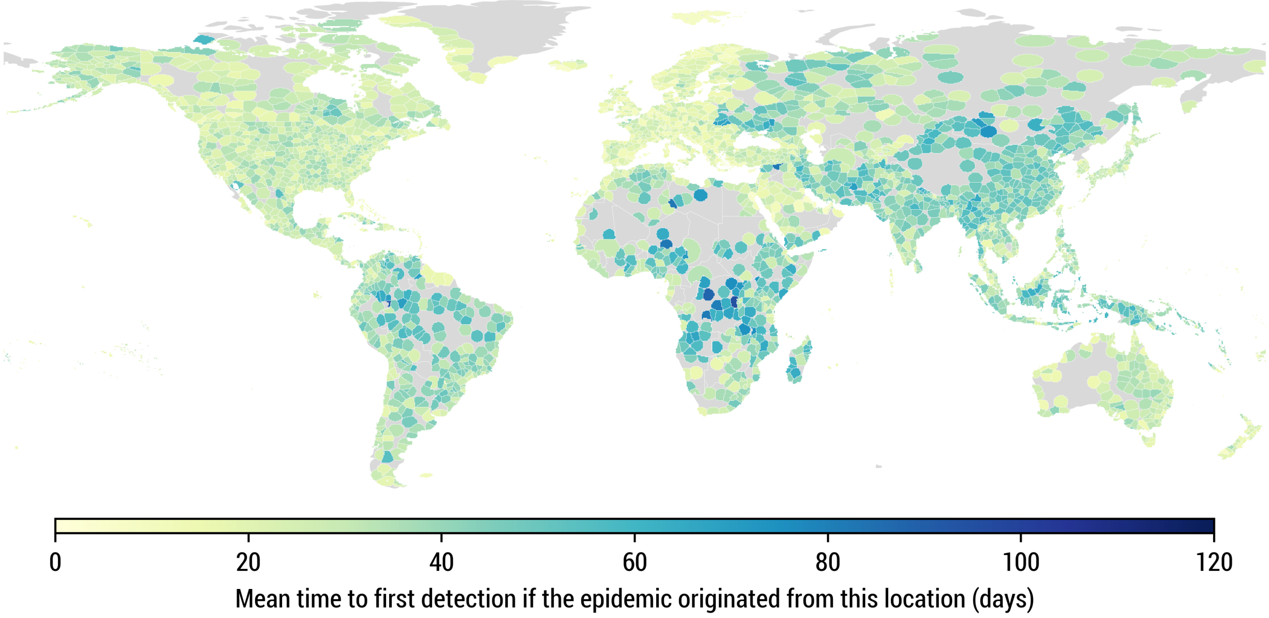

**Figure S6**: Mean time to first detection with the complete global surveillance network, with a sentinel at every airport. We use the same model parametrization as in Fig. 1.

where $\hat{T}_{\text{fd}}$ is the mean time to first detection with doubling time $\hat{T}_2$. This is illustrated in Fig. S9 for all potential origins, when changing the mean generation time or the reproduction number. The Pearson correlation coefficients are very high (>0.99).

To justify Eq. (24), let us rephrase more formally the argument introduced in the main text. If we neglect the stochastic fluctuations of the epidemic, the number of detectable individuals is approximately $D(t) \simeq D_0 e^{\lambda t}$, $t$ days after the beginning of the outbreak. Let us also assume that each day, a detectable individual has some constant probability $\omega$ to travel and be detected by the WWSN. In that case, we can approximate the probability of having a first detection on day $t$ as [29]

$$P(t_{\text{fd}} = t) \approx [1 - (1-\omega)^{D(t)}] \prod_{t'=1}^{t-1} (1-\omega)^{D(t')} \tag{26}$$

$$\approx \xi e^{\lambda t} \exp\left(-\frac{\xi}{\lambda} e^{\lambda t}\right) , \tag{27}$$

where $\xi = \omega D_0$, and the approximations hold when $\xi/\lambda \ll 1$. Since the probability per day of air-travel is very small in general (see Fig. S7), this approximation is almost always valid. We recognize a Gumbel distribution with mean

$$T_{\text{fd}} = \frac{1}{\lambda}\left[\ln\left(\frac{\lambda}{\xi}\right) - \gamma\right] , \tag{28}$$

where $\gamma$ is the Euler–Mascheroni constant. Rearranging the terms and using $\lambda = \ln 2/T_2$, we get

$$\frac{T_{\text{fd}}}{T_2} + \log_2 T_2 = \frac{1}{\ln 2}\left(\ln(\ln 2) - \ln \xi - \gamma\right) = \text{const.} , \tag{29}$$

which is identical to Eq. (24).

In Fig. S10, we show that the Gumbel distribution approximates well the distribution for the time to first detection, especially when the initial number of latent and infectious is high in Fig. S10(B). This is due to the fact that the

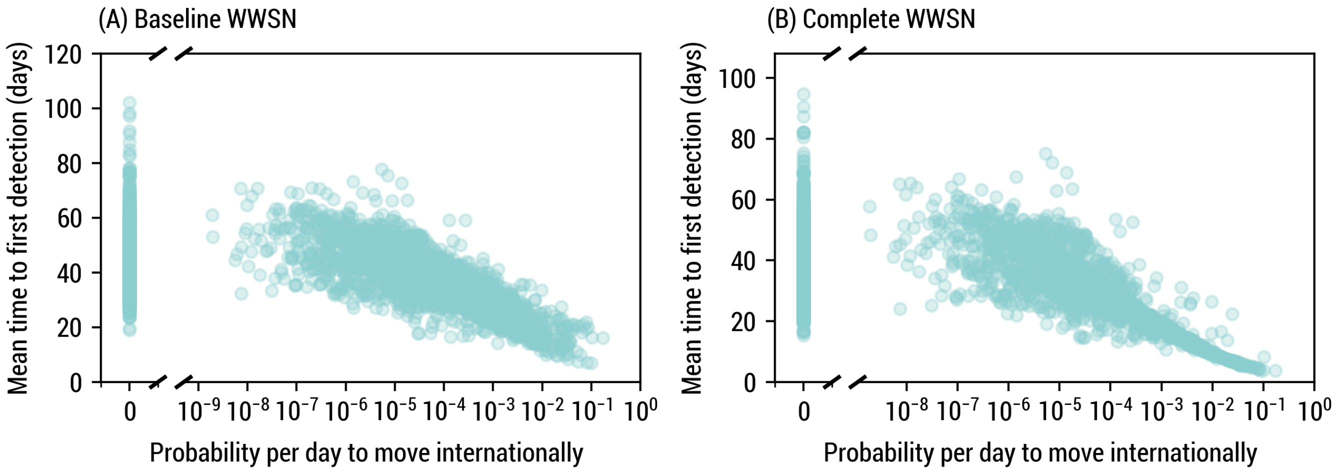

**Figure S7**: Mean time to first detection considering each of the 3200+ subpopulations as potential origin, against the probability per day to move internationally from each subpopulation. We use the same model parametrization as in Fig. 1. (A) We use the same baseline WWSN as in Fig. 1. The Pearson correlation coefficient is -0.810 (90% CI, -0.821 to -0.799; two-sided $P$ value <0.001) between the mean time to first detection and the logarithm of the probability per day to move internationally (for subpopulations where this probability is larger than zero, $n = 2509$). (B) We use the complete WWSN, with a sentinel at every airport. The Pearson correlation coefficient is -0.862 (90% CI, -0.870 to -0.853; two-sided $P$ value <0.001) between the mean time to first detection and the logarithm of the probability per day to move internationally (for subpopulations where this probability is larger than zero, $n = 2509$).

Gumbel approximation neglects the stochastic fluctuations of the epidemic, which are more important when starting with fewer exposed individuals.

To obtain the Gumbel approximations in Fig. S10, we calculate $\lambda$ and $T_{\mathrm{fd}}$ from our PGF framework, which allows us to fix $\xi$ in Eq. (28). While $\xi$ should not depend on $\lambda$, we do find small variations, which also explain the small variations of the approximate invariant quantity in Eq. (24) and in Fig. 3. This is because we treated the detectable individuals $D(t)$ as one homogeneous population with a fixed probability of moving and being detected $\omega$, while in fact it is a heterogeneous group of individuals distributed across a complex metapopulation network.

#### 2.2 Sensitivity analyses

Other than the reproduction number and the generation time that directly affect the growth rate, other aspects of the disease's natural history impact the time to first detection, but less significantly so. Here we vary the length of the post-infectious period—thereby changing the detectable period—and the shape of the secondary-infection distribution. In our framework, we do not directly fix the secondary-infection distribution, but rather the secondary-infection distribution per time step. To tune the variance, we replace the multivariate Poisson PGF term in the reaction phase (see Table S1) by the composition of a multinomial and a negative binomial PGF, namely

$$\exp\left[\sum_{\alpha'} \beta_{\alpha,\alpha'}(z_{\alpha'} - 1)\Delta t\right] \mapsto \left[1 + \frac{\Delta t}{\kappa}\sum_{\alpha'} \beta_{\alpha,\alpha'}(1 - z_{\alpha'})\right]^{-\kappa} . \tag{30}$$

This results in the number of secondary infections per time step being distributed according to a negative binomial [34], a generalization of the Poisson distribution. The composition with the multinomial PGF ensures contacts are made according to the age-structured contact matrix. Increasing the overdispersion parameter $\kappa$ reduces the variance while

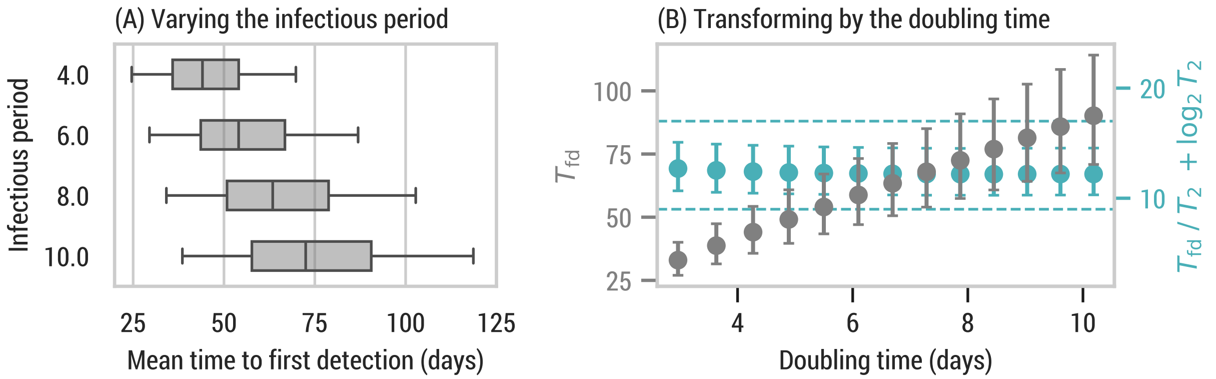

**Figure S8**: Impact on the mean time to first detection of varying the generation time via the infectious period instead of the latent period. We keep the detectable period fixed to 15 days by also changing the post-infectious detectable period as $T_{\text{post}} = T_{\text{det}} - T_{\text{inf}}$. Otherwise, we use the same WWSN and parametrization as in Fig. 1. All prediction intervals are obtained from $n = 3244$ subpopulations. (A) The center line of the box plot indicates the median, the box covers the interquartile range and the whiskers cover the 90% central prediction interval; the outliers outside the interval are not shown. (B) Circles indicate the median and the error bars cover the interquartile range. The dashed lines are there to guide the eyes. We vary the infectious period between 2 and 14 days, resulting in doubling times between 3 and 10.2 days.

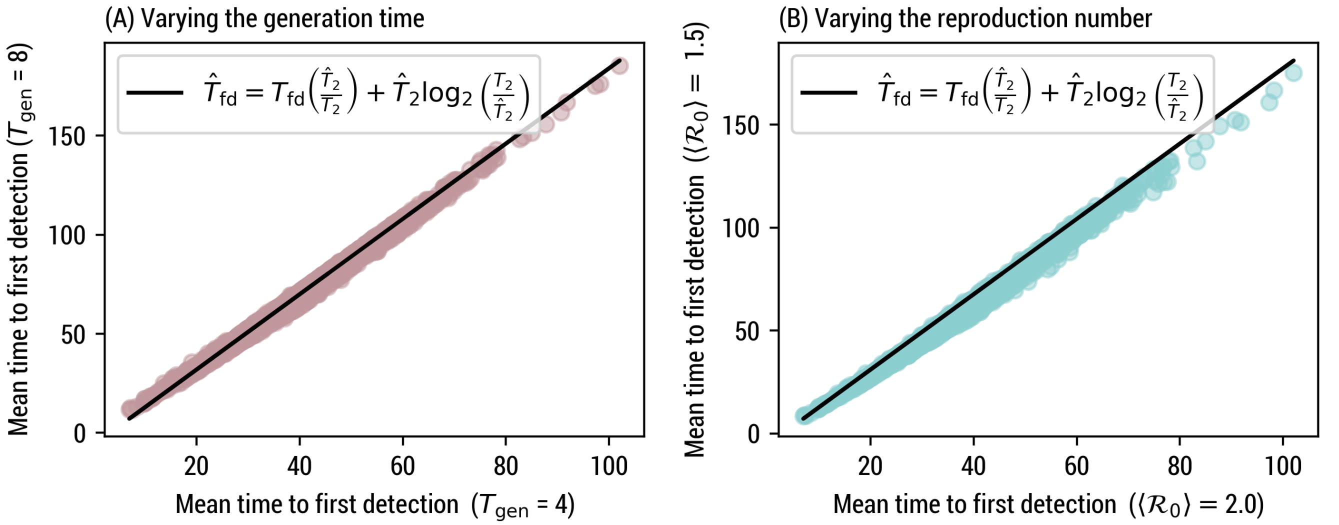

**Figure S9**: Linear transformation of the mean time to first detection for varying natural disease history. We use the same WWSN as in Fig. 1. The detection probability is 16% and the detectable period is 12.7 days. (A) The mean reproduction number is fixed $\langle \mathcal{R}_0 \rangle = 2$, and the mean generation time varies. The Pearson correlation coefficient is 0.997 with a two-sided $P$ value $< 0.001$ ($n = 3244$). (B) The mean generation time is fixed $T_{\text{gen}} = 4$, and the reproduction number varies. The Pearson correlation coefficient is 0.996 with a two-sided $P$ value $< 0.001$ ($n = 3244$).

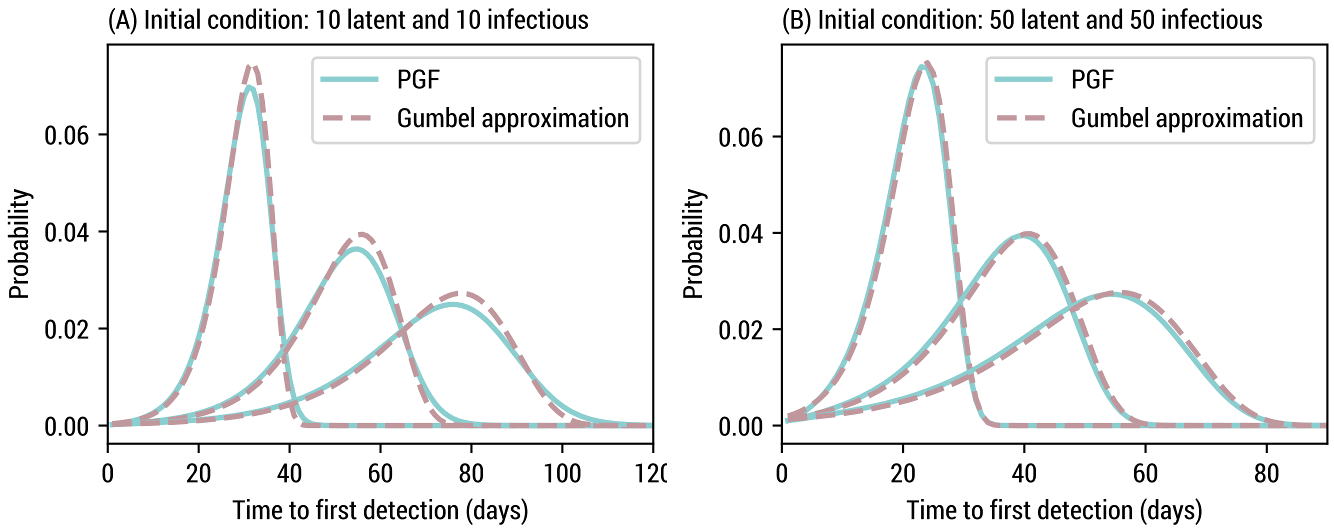

**Figure S10**: Approximation of the time to first detection using a Gumbel distribution. We use the same WWSN as in Fig. 1 and, unless specified, the same parametrization. We focus on an epidemic originating in São Paulo. We show the results for mean generation times of 4, 8, and 12 days. We consider two initial conditions: (A) 10 latent and 10 infectious individuals and (B) 50 latent and 50 infectious individuals.

**Table S3**: Statistics of the mean time to first detection for various post-infectious periods and overdispersion parameters for the secondary-infection distribution (per time step). The rest of the model parameters and the WWSN are the same as in Fig. 1. We report the median and the 5th and 95th percentile in parentheses ($n = 3244$).

| Overdispersion $\kappa$ | | 0.01 | 0.03 | 0.1 | $\infty$ |
|---|---|---|---|---|---|
| Infections caused by the top 20% | | 81.9% | 66.2% | 56.8% | 51.3% |
| Post-Infec. period | 5 | 39.6 (22.4,62.1) | 38.3 (21.4,60.7) | 37.9 (21.1,60.1) | 37.7 (21.0,59.9) |
| | 10 | 38.9 (21.7,61.4) | 37.7 (20.8,60.0) | 37.2 (20.4,59.4) | 37.0 (20.3,59.1) |
| | 20 | 38.5 (21.3,60.9) | 37.2 (20.4,59.5) | 36.8 (20.1,58.9) | 36.6 (20.0,58.7) |

reducing $\kappa$ increases the variance[1]; in the limit $\kappa \to \infty$, we recover the multivariate Poisson PGF.

Another way to communicate the overdispersion in epidemiology is to assess what portion of infections are caused by the top 20% of infectors [34]. We compile our results in Table S3 for various combinations of post-infectious period and overdispersion parameters. Overall, both have a limited impact on the distribution of $T_{\text{fd}}$ for all subpopulations. A broader secondary-infection distribution will mainly affect the beginning of an epidemic—once the outbreak is large enough, the stochastic fluctuations are averaged out. Similar results were obtained in Ref. [6]. The small impact of changing the detectable period (through the post-infectious period) is due to the fact that the number of infectious grows exponentially at the beginning of an epidemic. Consequently, unless the growth rate of the disease is very small, detection at airports should be predominantly caused by newly infectious individuals traveling.

Variation of the air-travel patterns also do not lead to large variations of the global statistics of $T_{\text{fd}}$ for all locations, as illustrated in Fig. S11. The median of the distribution for $T_{\text{fd}}$ varies from 36 days in the summer to 38 days in the Fall, and the 90% prediction intervals remain relatively stable.

[1] It is important to distinguish the overdispersion parameter $\kappa$ given here, associated to the secondary-infection distribution per time step of duration $\Delta t$, versus what one would expect using a negative binomial for the secondary-infection distribution over the whole infectious period.

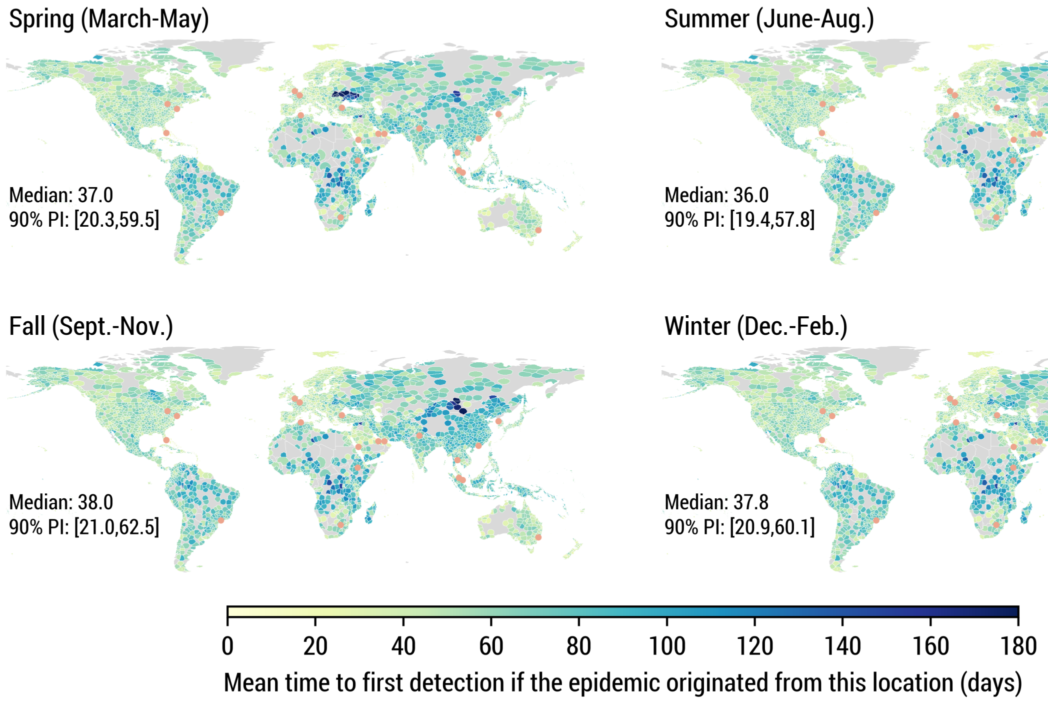

**Figure S11**: Mean time to first detection using different air-travel mobility networks associated with different seasons. We use the same WWSN and model parametrization as in Fig. 1. Statistics reported are based on $n = 3244$ subpopulations.

#### 2.3 Optimization of the time to first detection

In the Methods section of the main text, we introduce a greedy optimization scheme to minimize the mean time to first detection, averaged over all subpopulations as a potential source

$$\Phi(\mathcal{S}) = \sum_{l} P(l) T_{\mathrm{fd}}(\mathcal{S}, l) \ , \tag{31}$$

with probability $P(l)$ of being the source. In the second step of the optimization procedure, it requires the evaluation of $\Phi(\mathcal{S} \cup \{\nu\})$ for each airport $\nu \notin \mathcal{S}$ to assess the reduction in time to first detection associated with adding $\nu$ as a sentinel. However, evaluating $\Phi(\mathcal{S} \cup \{\nu\})$ for each new potential sentinel is computationally expensive.

Instead, we use an approximation for each $T_{\mathrm{fd}}(\mathcal{S}, l)$ hinging on two assumptions. First, we assume the full distribution of the time to first detection from each source subpopulation $l$ is well-described by a Gumbel distribution—which is validated in Fig. S10. Second, we assume that the detection at all sentinels $\nu \in \mathcal{S}$ are independent processes. From these assumptions, we can leverage the following identity [29]

$$\exp\left(-\lambda T_{\mathrm{fd}}(\mathcal{S}, l)\right) = \sum_{\nu \in \mathcal{S}} \exp\left(-\lambda T_{\mathrm{fd}}(\{\nu\}, l)\right) \ . \tag{32}$$

This allows us to efficiently estimate $T_{\mathrm{fd}}(\mathcal{S}, l)$ from the individual $T_{\mathrm{fd}}(\{\nu\}, l)$ for all $\nu \in \mathcal{S}$. Figure S12 validates the accuracy of the approximation.

##### Submodularity proof

In the context of the approximation provided by Eq. (32), we want to show that $-\Phi(\mathcal{S})$ is a monotone submodular set function. Since $-\Phi(\mathcal{S})$ is a positive linear combination of $-T_{\mathrm{fd}}(\mathcal{S}, l)$ for each potential source $l$, it suffices to show that $-T_{\mathrm{fd}}(\mathcal{S}, l)$ is a monotone submodular set function for all $l$ [35].

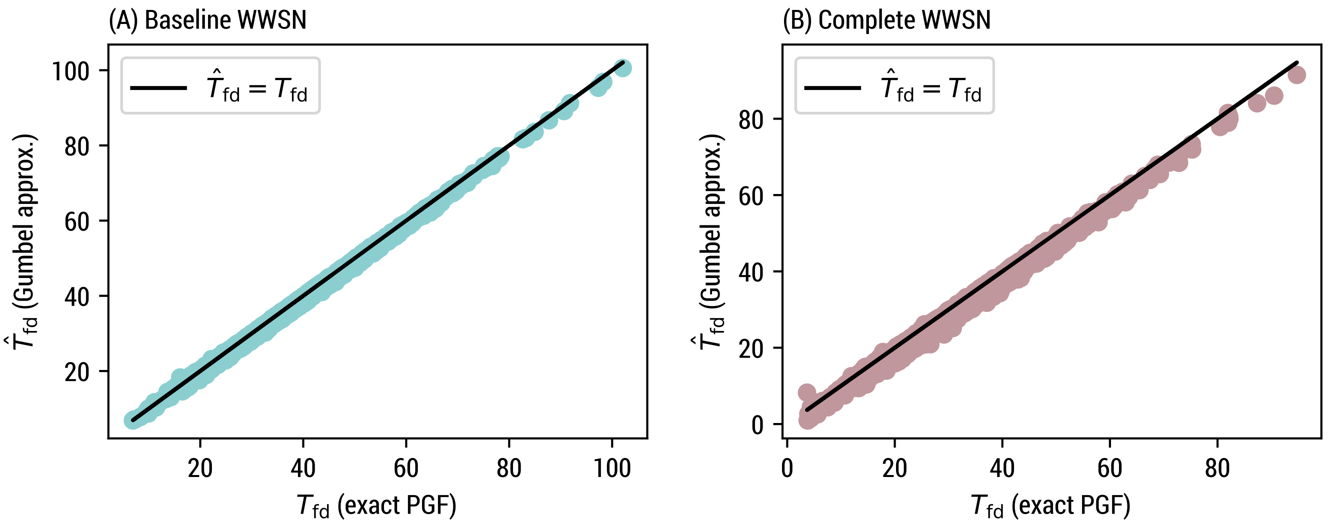

**Figure S12**: Comparison of the exact PGF calculation and the Gumbel approximation provided by Eq. (32) for the mean time to first detection, considering each of the 3200+ subpopulations as the origin. We use the same model parametrization as in Fig. 1. (A) We use the baseline WWSN. The Pearson correlation coefficient is > 0.999 with a two-sided $P$ value < 0.001 ($n = 3244$). (B) We use the complete WWSN. The Pearson correlation coefficient is 0.997 with a two-sided $P$ value < 0.001 ($n = 3244$).

We define the set of all potential sentinels as $\Omega$. Let us also simplify the notation by writing $\tau(B) \equiv T_{\text{fd}}(B, l)$ and $\tau(b) \equiv T_{\text{fd}}(\{b\}, l)$ for the mean time to first detection for every set of sentinels $B \subseteq \Omega$ and every sentinel $b$ respectively[2]. Note that $\tau : 2^\Omega \to \mathbb{R}^+$ is a set function, where $2^\Omega$ is the set of all subsets of $\Omega$

**Lemma 1.** *$\tau$ and $-\tau$ are monotone functions, i.e., $\tau(B) \leq \tau(A) \iff -\tau(B) \geq -\tau(A)$ for $A \subseteq B$.*

*Proof.* Let us define $C \equiv A \cap B$. From Eq. (32), we have

$$e^{-\lambda\tau(B)} = e^{-\lambda\tau(A)} + e^{-\lambda\tau(C)} \geq e^{-\lambda\tau(A)} \,, \tag{33}$$

where $\lambda > 0$. Taking the logarithm on both sides and dividing by $\lambda$, this implies $-\tau(B) \geq -\tau(A)$. □

**Definition 1.** *Let $\Omega$ be a finite set and $f : 2^\Omega \to \mathbb{R}$ a set function. The function $f$ is submodular if for two subsets $A, B \subseteq \Omega$ with $A \subseteq B$ and $e \in \Omega \setminus B$, we have $f(A \cup \{e\}) - f(A) \geq f(B \cup \{e\}) - f(B)$ [35].*

**Theorem 1.** *The function $-\tau$ is monotone and submodular.*

*Proof.* The function $-\tau$ is monotone from lemma 1. Let us consider two sets $A$ and $B$ such that $A \subseteq B \subseteq \Omega$, $C \equiv B \setminus A$, and an element $e \in \Omega \setminus B$. Using Eq. (32), we have

$$\begin{aligned} e^{-\lambda\tau(B\cup\{e\})} &= e^{-\lambda\tau(B)} + e^{-\lambda\tau(e)} \,, \\ &= e^{-\lambda\tau(A)} + e^{-\lambda\tau(C)} + e^{-\lambda\tau(e)} \,, \end{aligned}$$

where $\lambda > 0$. Dividing both sides by $e^{-\lambda\tau(B)}$ and applying again Eq. (32), we find

$$e^{-\lambda[\tau(B\cup\{e\})-\tau(B)]} = \frac{e^{-\lambda\tau(A\cup\{e\})} + e^{-\lambda\tau(C)}}{e^{-\lambda\tau(A)} + e^{-\lambda\tau(C)}} \leq e^{-\lambda[\tau(A\cup\{e\})-\tau(A)]} \,, \tag{34}$$

[2] The ensuing results are general for all epidemic source $l$ and therefore we drop the index to simplify the notation.

where the inequality holds if and only if $\tau(A \cup \{e\}) \leq \tau(A)$, which is true from lemma 1, but also more intuitively because adding sentinels can only reduce the time to first detection. Taking the logarithm on both sides of Eq. (34) and dividing by $\lambda$, we arrive at

$$-\tau(B \cup \{e\}) + \tau(B) \leq -\tau(A \cup \{e\}) + \tau(A) \; . \tag{35}$$

From definition 1, this implies that $-\tau$ is a submodular set function. □

### 3 Retrospective counterfactual scenarios

#### 3.1 SARS-CoV-2 Alpha variant emergence

In the main text, we present the results of a counterfactual scenario where we would have had a global WWSN to track the international dissemination of the SARS-CoV-2 Alpha (B.1.1.7) variant. We utilize air-travel data from September 2020 to November 2020, along with the baseline WWSN of 20 sentinels (see Table S4).

This variant was first identified by health authorities in the United Kingdom and retrospective analyses trace the first identified case in Kent, South East England, on September 20, 2020 [36]. According to data published by the United Kingdom government [37], the effective reproduction number for SARS-CoV-2 between September 11 and October 30, 2020, was between 1.1 and 1.4 (90% CI) in both London and South East England. The Alpha variant was found to be more transmissible, with an estimated increased reproduction number ranging from 40 to 100% [36, 38, 39]. In this counterfactual study, we considered a value of $\mathcal{R}_{\text{eff}}^{\text{alpha}} = 1.7$ for the Alpha variant, on the lower side of available estimates, equivalent to having an effective reproduction number of $\mathcal{R}_{\text{eff}}^{\text{ws}} = 1.1$ for the wild strain and an increased transmissibility of 55%. Since approximately 5% of positive cases were sequenced at the source [36], we considered an initial cluster of 20 infectious and 20 latent individuals on September 15. The generation time is kept fixed at 6.5 days, with a latency period of 4.5 days.

Distributions for the time to first detection in Fig. 5A are calculated using the PGF methodology and are not influenced by the wastewater sampling scheme. The geolocalization of the source (Fig. 5B) and the parameter inference (Fig. 5C), however, require us to transform distributions for the number detections to acccount for wastewater pool sampling at sentinel sites.

##### Distribution for the cumulative number of detections by a WWSN

Our PGF methodology allows us to estimate $P(d_t = d)$, the distribution for the cumulative number of detections at time $t$ by the WWSN. However, as defined, all detectable agents traveling through a screened route could contribute to $d_t$ *independently*. While this can be a reasonable assumption for individual testing, this is not the case for wastewater sampling, where it is not usually feasible to identify precisely the number of detectable individuals in an aircraft—we only get a binary *yes* or *no* answer to whether or not the pathogen was detected. Depending on how the testing is performed—on individual aircraft, or pooled sampling of multiple aircraft at a triturator—we would obtain different results for the cumulative number of detections.

We assume the pooled testing of all aircraft within a day at each sentinel airport. To take into account the potential copresence on the same day of detectable individuals traveling to the same destination, we need to define the probability $P(\tilde{d}|d)$, where $\tilde{d}$ represents the cumulative "wastewater" detections and $d$ the cumulative "individual" detections we would obtain if we were able to detect each individual independently. Then we get the adjusted distribution

$$P(\tilde{d}) = \sum_{d} P(\tilde{d}|d) P(d) \; . \tag{36}$$

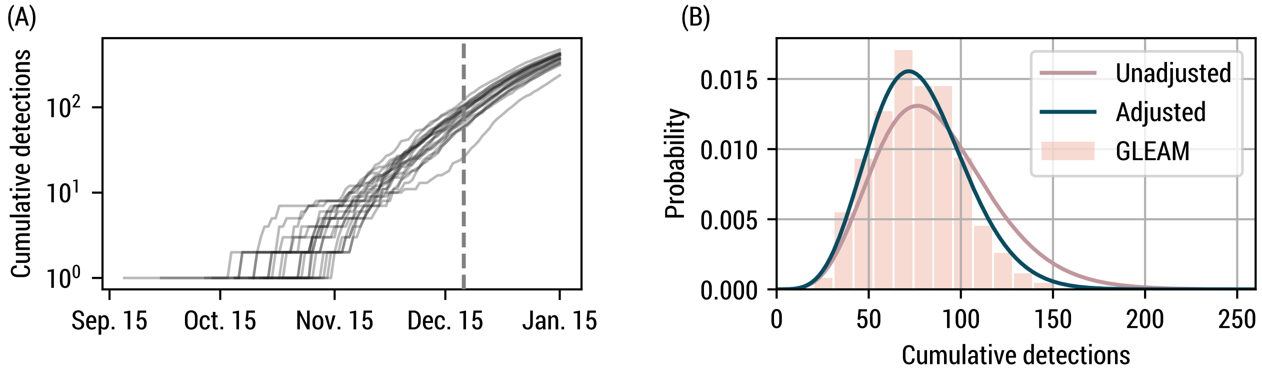

**Figure S13**: Cumulative number of wastewater detections for the Alpha variant counterfactual study. (A) Example of 20 detection time series from GLEAM simulations. The vertical dashed line indicates December 20, 2020. (B) Distribution for the cumulative number of detections on December 20, 2020, from 1250 GLEAM simulations and the PGF methodology, with and without adjustment for wastewater pool sampling.

We evaluate $P(\tilde{d}|d)$ by first characterizing $q_{\nu,t}$, the relative probability an individual detection happens at sentinel $\nu$ and at time $t$. In general, $q_{\nu,t} \propto \langle d^*_{\nu,t}\rangle$, where $\langle d^*_{\nu,t}\rangle$ is the mean number of individual detections at sentinel $\nu$ and *incident* on time $t$—not the cumulative detections. The ratio $\langle d^*_{\nu,t}\rangle/\langle d^*_{\nu',t}\rangle$ is very stable in the early phase of an outbreak for any pair of airports $(\nu,\nu')$. Therefore, $\langle d^*_{\nu,t}\rangle \sim A(\nu)e^{\lambda t}$, where $\lambda$ is the growth rate at the source and $A(\nu)$ is the relative propensity of detection at each sentinel airports in the early phase. After normalization, we encapsulate all probabilities in a vector $\boldsymbol{q}$. Secondly, we assume that the number of *incident* detections for each sentinel and day $d^*_{\nu,t}$ ($\boldsymbol{d}^*$ in vector format) is distributed according to a multinomial $P(\boldsymbol{d}^*)$ with parameters $d$ (the sum of all $\boldsymbol{d}^*$) and $\boldsymbol{q}$. The total number of *wasterwater* detections corresponds to the number of nonzero counts in $\boldsymbol{d}^*$, and thus $P(\tilde{d}|d)$ is obtained by summing $P(\boldsymbol{d}^*)$ over all configurations such that there are a total of $\tilde{d}$ nonzero counts. While this is technically a difficult combinatorial task, this can be carried out efficiently numerically (see Ref. [40]).

In Fig. S13(A), we show examples of time series for the cumulative number of detections at all sentinels of the WWSN generated by GLEAM. In Fig. S13(B), we illustrate the importance of accounting for wastewater pooled sampling when the number of detections is sufficiently large; the number of wastewater detections is effectively reduced due to the copresence of detectable individuals transiting through as sentinel during the same day.

##### Geolocalization of the source

Even without information about which flight paths led to detections, it is possible to recover information about the location of the source of an epidemic. Indeed, we can construct a likelihood $P(\tilde{\boldsymbol{d}}|\text{source} = l) \equiv P(\tilde{\boldsymbol{d}}|l)$ for the probability to have observed the wastewater detections $\tilde{\boldsymbol{d}} = (\tilde{d}_\nu)_{\nu\in\mathcal{S}}$ at the sentinel airports.

First, let us ignore wastewater pool sampling and consider individual detections $\boldsymbol{d} = (d_\nu)_{\nu\in\mathcal{S}}$ and the associated likelihood function $P(\boldsymbol{d}|l)$. Then $P(\boldsymbol{d}|l)$ can be modeled by a multinomial distribution with parameter $d = \sum_\nu d_\nu$ and probability vector $\boldsymbol{q} = (q_\nu)_{\nu\in\mathcal{S}}$, where $q_\nu \propto \langle d_\nu\rangle$, the expected number of detections at sentinel $\nu$ when we have $d$ detections. In practice, we approximate $q_\nu$ by the expected number of detections at each sentinel when we have a first detection.

To account for wastewater pool sampling, we transform the likelihood as

$$P(\tilde{\boldsymbol{d}}|l) = \sum_{\boldsymbol{d}} P(\tilde{\boldsymbol{d}}, \boldsymbol{d}|l) = \sum_{\boldsymbol{d}} P(\tilde{\boldsymbol{d}}|\boldsymbol{d}, l)P(\boldsymbol{d}|l) \ . \tag{37}$$

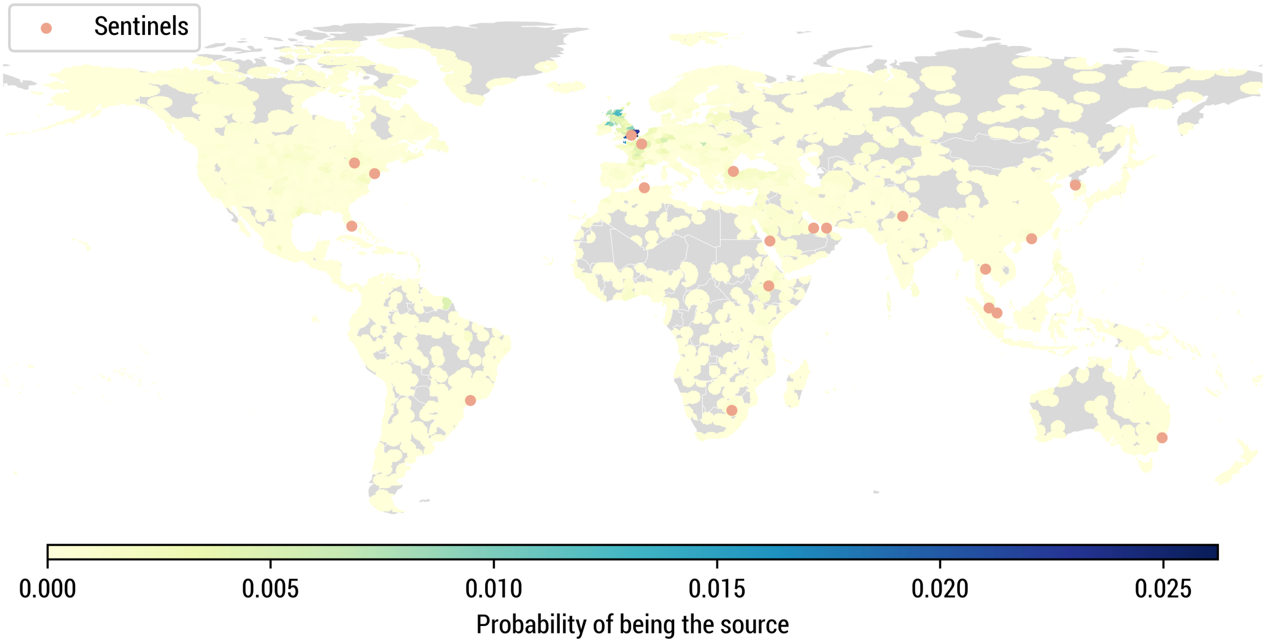

**Figure S14**: Geolocalization of the source for the SARS-CoV-2 Alpha variant counterfactual scenario. We show the posterior distribution over subpopulations averaged over 1250 GLEAM simulations, when $\geq$ 10 cumulative wastewater detections have occurred. We use the same model parametrization as in Fig. 5 for the geolocalization.

Assuming that $P(\tilde{\boldsymbol{d}}|\boldsymbol{d}, l)$ is concentrated around its mean, we can use the following approximation

$$P(\tilde{\boldsymbol{d}}|l) \approx \sum_{\boldsymbol{d}} \delta_{\boldsymbol{d},\boldsymbol{b}} P(\boldsymbol{d}|l) = P(\boldsymbol{b}|l) \;, \tag{38}$$

where $\boldsymbol{b}$ is the expected number of "individual detections" given the observed pooled wastewater detections $\tilde{\boldsymbol{d}}$, rounded to the nearest integer.

Using the likelihood in Eq. (38), we compute the following posterior distribution for the source

$$P(l|\tilde{\boldsymbol{d}}) \propto P(\tilde{\boldsymbol{d}}|l)P(l) \;, \tag{39}$$

where $P(l)$ is a prior distribution on the source location. In this work, we consider a uniform prior $P(l) = \text{const}$. In Fig. S14, we show this posterior distribution for at least 10 wastewater detections, averaged over 1250 simulations. We see that most of the posterior density is concentrated in Europe, and especially in the United Kingdom. The same simulations and posterior distributions are used to assess the geolocalization capacities of a WWSN in Fig. 5, as detections accumulates at the sentinels.

#### Characterization of the growth dynamics

The time series of cumulative detections can also be utilized to estimate key epidemic parameters. For generic parameter inference, we use the following approach. Given the observed cumulative number of wastewater detections $\tilde{d}$ and $\tilde{d}'$ at different time $t$ and $t'$, we calculate the posterior distribution over $\boldsymbol{\theta}$ (the parameters) using

$$P(\boldsymbol{\theta}|\tilde{d}', \tilde{d}) \propto P(\tilde{d}', \tilde{d}|\boldsymbol{\theta})P(\boldsymbol{\theta}) \;. \tag{40}$$

To compute more efficiently the joint likelihood $P(\tilde{d}', \tilde{d}|\boldsymbol{\theta})$, we evaluate the joint cumulants from the CGF and employ the method of moments with a negative multinomial distribution.

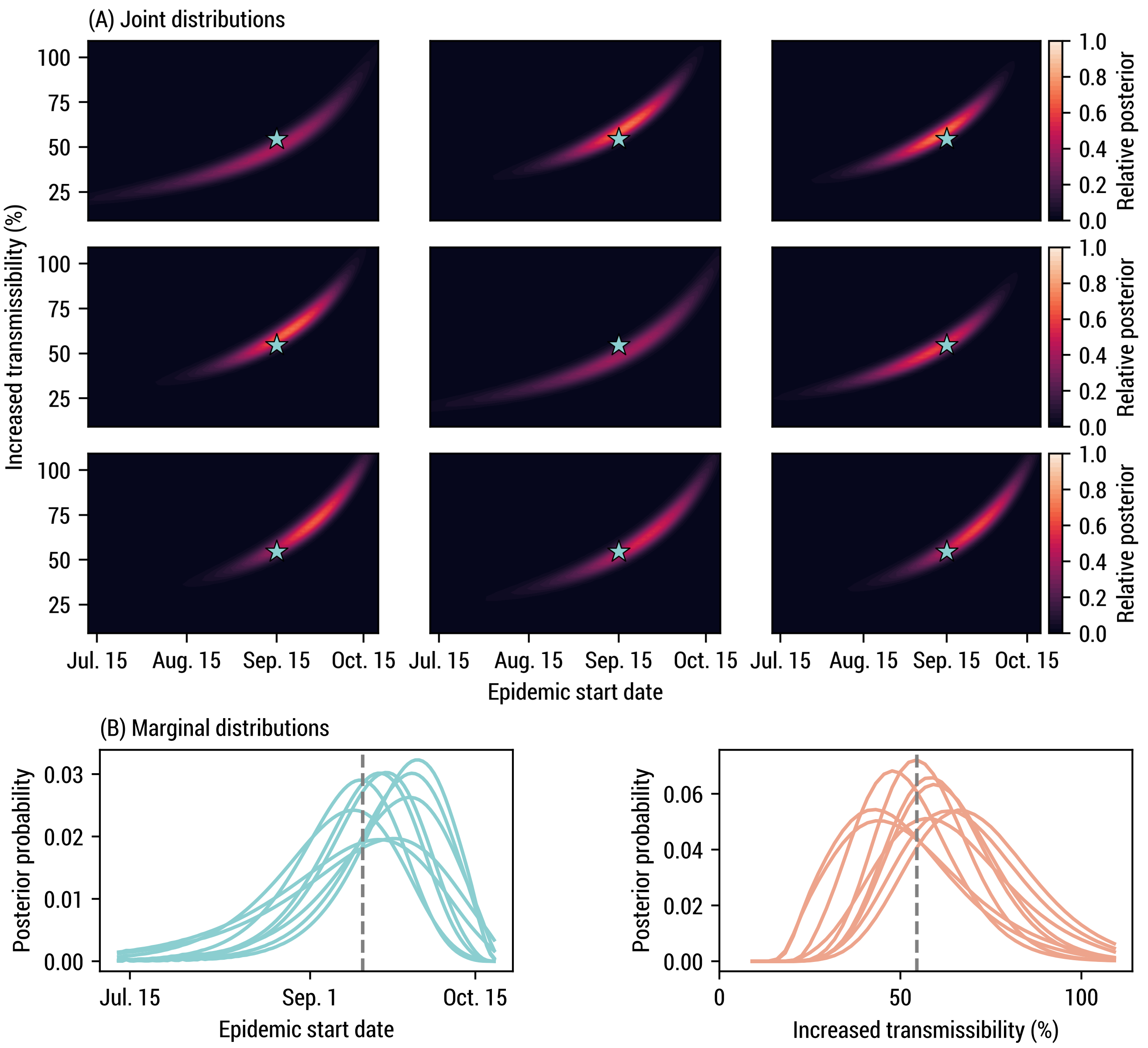

**Figure S15**: Examples of posterior distributions obtained for a subset of detection time series ($n = 9$) used to construct the average posterior distribution in Fig. 5C. (A) Joint posterior distributions. The blue star represent the ground truth (55% increased transmissibility and September 15, 2020 as the start date). (B) Marginal distributions, integrating over each axis. The vertical dashed line indicate the ground truth.

In Fig. 5C of the main text, we jointly infer the epidemic start date and the increased transmissibility of the Alpha variant with respect to the SARS-CoV-2 wild strain. We use a flat prior $P(\boldsymbol{\theta}) = \text{const.}$, between 9% and 109% for the increased transmissibility ($\mathcal{R}_{\text{eff}}^{\text{alpha}} \in [1.2, 2.3]$) and between July 12 and October 20, 2020 for the starting date (cluster of 20 latent and 20 infectious). In Fig. S15, we also illustrate a subset of the posterior distributions from individual time series.

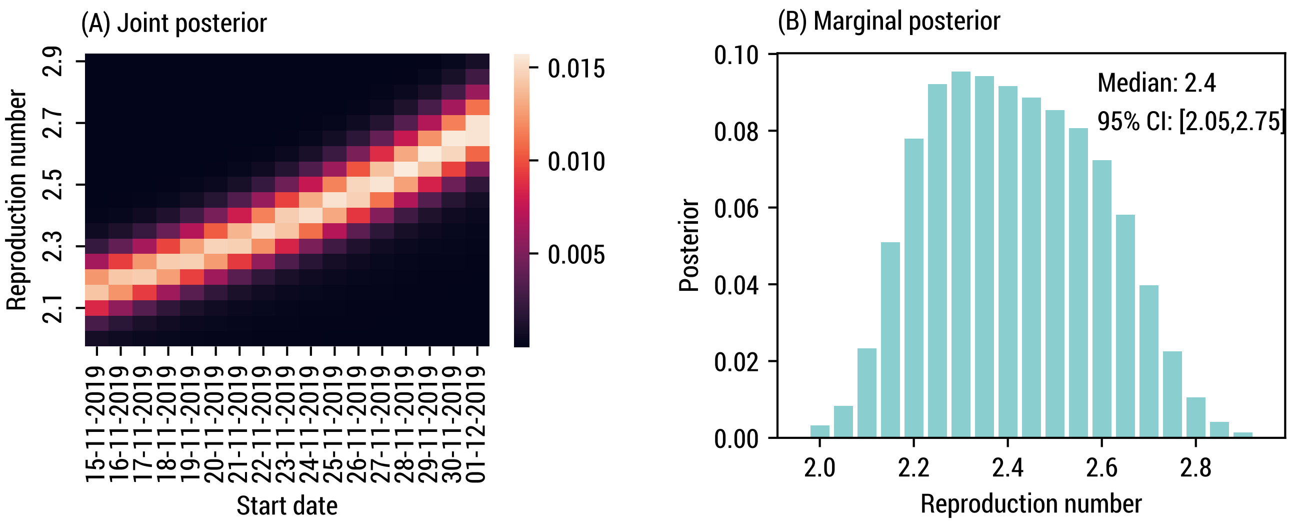

**Figure S16**: Calibration on international importations for the emergence of SARS-CoV-2 (wild strain). We assume an initial cluster of 10 infectious and 10 latent individuals, a generation time of 6.5 days and a detectable period of 12.7 days. (A-B) Posterior distributions given 69 international COVID-19 cases with a travel history and an arrival date before or on Jan. 23, 2020.

#### 3.2 SARS-CoV-2 (wild strain) emergence

To assess the potential effectiveness of a WWSN in early detection and response to a pandemic situation, we analyze a second hypothetical scenario: the operation of a WWSN prior to the onset of the COVID-19 pandemic. This counterfactual study utilizes air-travel data from December 2018 to February 2019, along with the baseline WWSN of 20 sentinels (see Table S4).

###### Calibration on importations

Considering the high uncertainty at the beginning of the pandemic, we first calibrate our model to available data. To do so, we use the same approach as in Refs. [5, 6], i.e., we calibrate our model on the number of international importations until January 23rd, 2020, when travel restrictions were imposed.

Only a fraction of importations are identified at the destination, notably because of asymptomatic individuals—we considered a 40% rate of asymptomatic individuals [41]—, but also because of different levels of capacity for detection. To account for the heterogeneity in case detection, we stratify countries into three groups based on the second component of the Global Health Security Index: high ($\geq$80th percentile), low ($\leq$20th percentile), and medium (the rest) surveillance capacity. We then use the estimates provided in Ref. [42] to assign a probability of detection relative to Singapore, which has had strong epidemiological surveillance in past infectious disease outbreaks including the COVID-19 pandemic. We used a 60% probability of detection for symptomatic individuals in Singapore, and then countries in the high, medium, and low categories were assigned relative capacities corresponding to 40%, 37%, and 11% compared to Singapore.

To calibrate our model, we compute the distribution $P(E|\boldsymbol{\theta})$ for the number of importations $E$ given the parameters $\boldsymbol{\theta}$ of the model—in the present case, $\boldsymbol{\theta}$ corresponds to the start date and the basic reproduction number. This likelihood is used to evaluate the posterior distribution $P(\boldsymbol{\theta}|E) \propto P(E|\boldsymbol{\theta})$. We use a uniform prior between November 15th and December 1st, 2019 for the starting date with an initial cluster of 10 infectious and 10 latent individuals in Wuhan, consistent with estimates placing the index case mid-October to mid-November [43]. We also consider a

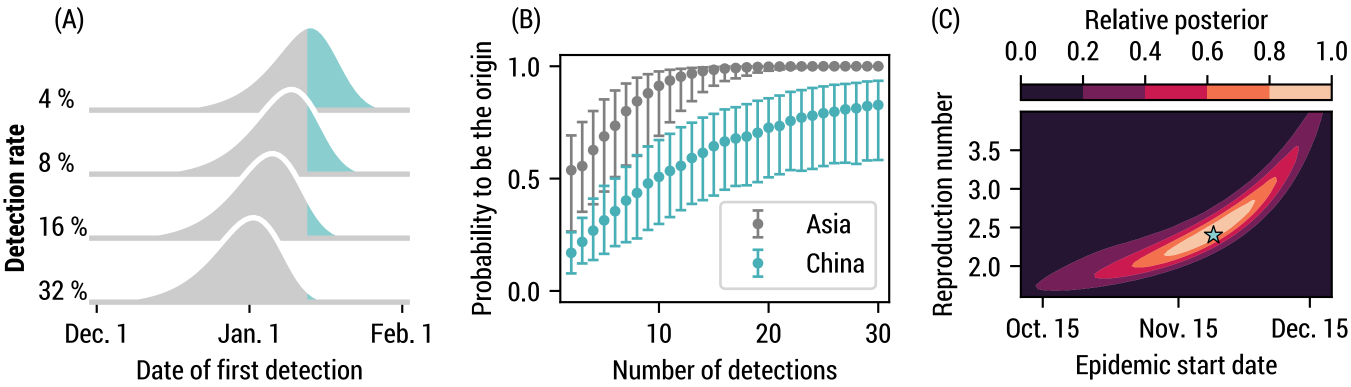

**Figure S17**: A global WWSN would provide an early warning system for international spreading and timely inferential capabilities. We consider a counterfactual scenario of the emergence of COVID-19 where a global WWSN would have been available. We use the baseline surveillance system consisting of 20 sentinels (see Table S4). We calibrate our model on international importations with an arrival date on or before Jan. 23 2020. (A) Distributions for the time to first detection with varying detection rates. The blue portion corresponds to dates after the first international case was reported in Thailand on January 13 [47]. (B)-(C) Inference experiment using data generated by the mechanistic GLEAM model with $\mathcal{R}_0 = 2.4$ and a start date of November 23 for the initial cluster. (B) Geolocalization of the source as more detections cumulate. We compute the posterior distribution for the origin of the epidemic based on the detection counts at each sentinel. Geolocalization of the source as more detections cumulate. We compute the posterior distribution for the origin of the epidemic based on the detection counts at each sentinel. The markers indicate the median posterior value and the whiskers the interquartile range obtained from 1200 detection time series. (C) Joint posterior distribution on $\mathcal{R}_0$ and the start date, averaged over 59 detection time series. The blue star indicates the ground truth for the simulation experiment.

uniform prior distribution on $\mathcal{R}_0$ between 2 and 2.9. The generation time is kept fixed at 6.5 days, with a latency period of 4.5 days.

In Fig. S16(A), we show the posterior distribution obtained for $\mathcal{R}_0$ and the starting date using as evidence 69 identified importations by Jan. 23, 2020 (see Table S1 in the Supplementary Material of Ref. [5]). Summing the joint posterior over the plausible starting date, we obtain the marginal posterior distribution on $\mathcal{R}_0$ in Fig. S16(B), with a median of 2.4 (95% CI, 2.05–2.75). The resulting median doubling time is 4.05 days (95% CI, 3.35—5.15), broadly consistent with other estimates [5, 6, 44–46].

##### Results

In Fig. S17, we present similar results as in Fig. 5, but for the emergence of SARS-CoV-2 in Wuhan, demonstrating the robust early warning and situational awareness capacities of a global WWSN.

We use the joint posterior of Fig. S16(A) to compute the posterior predictive distribution for the time to first detection in Fig. S17(A). We find that even with a low detection rate (4%), there is a 54% probability that an international case outside China would have been detected earlier than the first reported case in Thailand (January 13th), with a median time to first detection on January 12th. With a 16% detection rate—more in line with estimates for a triturator sampling scheme capturing all international inbound flights—the probability to have detection prior to January 13th is 91% and the median time to first detection is on January 4th.

This hypothetical scenario presupposes the capability of detecting SARS-CoV-2 in aircraft wastewater. The key point, however, is that an operational WWSN has the potential to significantly expedite the detection of international pathogen introductions. Any reduction in the time taken to sequence the genome of emerging pathogens would directly correlate to earlier detections. Furthermore, frozen samples can be stored and analyzed in the future.

Similarly to the Alpha variant retrospective study, we use synthetic time series data from GLEAM to illustrate how we can evaluate the source and key epidemic parameters. We fix the basic reproduction number to $\mathcal{R}_0 = 2.4$ and an initial cluster of 10 infectious and 10 latent individuals on November 23, 2019, based on the joint posterior in Fig. S16(A). We obtain the posterior distribution over subpopulations as a function of the number of wastewater detections and aggregate the posterior distribution at the continent and country level in Fig. S17(C). We find reliable geolocalization capacities at the country level after about 10 detections, with a median posterior probablity higher than 50%.

We also infer the basic reproduction number and the epidemic start date using the cumulative number of detections up until January 16 as the first observation $\tilde{d}$, and the cumulative number of detections between January 16 and January 23 as the second observation $\tilde{d}'$. We use a flat prior $P(\boldsymbol{\theta}) = \text{const.}$, between 1.5 and 4 for $\mathcal{R}_0$ and between October 5th and December 20th for the starting date (cluster of 10 latent and 10 infectious). The posterior distribution averaged over 59 time series is illustrated in Fig. S17(C). This average posterior indicates a median basic reproduction number of 2.4 (90% CI, 1.75–3.55), and a median starting date on November 20th (90% CI, Oct. 17–Dec. 14), illustrating the timely analytics a global WWSN could provide about the growh dynamics of emerging outbreaks.

#### 4 Airport table for the sentinel surveillance system

We report in Table S4 the list of sentinel airports for the baseline WWSN and the different optimization schemes (volume, entropy, greedy), when considering all subpopulations as equiprobable sources of an epidemic. Each airport is identified by its IATA code. We additionally list the sentinels obtained by the greedy approach in the context of targeted optimization, assuming all subpopulations within a specific continent as equiprobable sources of an epidemic, but all subpopulations outside the continent are ignored.

**Table S4**: Ordered list of sentinel airports (most important first) identified by the different optimization schemes. Each airport is identified by its IATA code [48]. We use the aggregated global air-travel network from September 2022 to August 2023. We also list the (unordered) baseline sentinels as determined in the main text.

| Global | | | | Targeted (greedy) | | | | | |
|---|---|---|---|---|---|---|---|---|---|
| baseline | volume | entropy | greedy | Africa | Asia | Europe | N. America | Oceania | S. America |
| ADD | DXB | FRA | LHR | DXB | DXB | IST | LHR | AKL | MIA |
| JNB | LHR | CDG | DXB | CDG | SIN | AMS | CUN | SIN | PTY |
| ALG | CDG | AMS | CDG | ADD | ICN | STN | YYZ | BNE | SCL |
| DXB | IST | LHR | SIN | IST | BKK | AYT | CDG | LAX | MAD |
| DOH | AMS | IST | IST | JNB | IST | FRA | MIA | HNL | LIS |
| JED | SIN | MUC | FRA | LIS | HKG | LGW | YVR | SYD | LIM |
| LHR | FRA | BRU | MIA | JED | JED | CPH | FRA | DPS | GRU |
| CDG | ICN | DXB | ICN | NBO | KUL | DXB | LAX | NAN | BOG |
| IST | MAD | VIE | AMS | LHR | TPE | DUB | MEX | MNL | AEP |
| JFK | DOH | DOH | CUN | MRS | DOH | VIE | AMS | SFO | EZE |
| YYZ | BKK | FCO | MAD | DOH | LHR | LHR | CPH | NRT | FLL |
| MIA | LGW | MXP | BKK | RUN | CAI | TLV | ICN | DXB | JFK |
| ICN | BCN | MAN | LAX | CAI | FRA | CDG | IAH | CDG | MEX |
| BKK | DUB | YYZ | DOH | BRU | DMK | EVN | PVR | KUL | CDG |
| DEL | HKG | STN | HKG | FRA | SHJ | MUC | PUJ | HKG | CUN |
| SIN | JFK | ZRH | YYZ | AMS | NRT | BCN | SJD | MEL | LHR |
| HKG | KUL | JFK | JED | EBB | SVO | TAS | FLL | ICN | MCO |
| KUL | FCO | LGW | LIS | CMN | KWI | ARN | YUL | LHR | SDQ |
| SYD | LIS | IAD | BNE | MXP | MNL | LTN | DFW | NOU | MVD |
| GRU | TPE | JNB | CPH | TUN | SGN | MAN | NRT | DOH | AMS |
| – | VIE | MAD | PTY | LYS | AUH | FCO | JFK | GUM | FRA |
| – | MUC | PRG | AYT | ABJ | MED | OSS | MNL | POM | HAV |
| – | ORY | DUS | TPE | MAD | CGK | PMI | YYC | BKK | PUJ |
| – | JED | ADD | KUL | BOM | CDG | DUS | PTY | CHC | FCO |
| – | YYZ | WAW | ADD | DZA | MCT | MAD | EWR | CNS | IAH |
| – | CPH | ORD | LGW | LPA | DEL | GYD | MBJ | YVR | YYZ |
| – | STN | YUL | NRT | KGL | KIX | ZRH | GDL | DEL | ATL |
| – | ZRH | EDI | JFK | ACC | DUS | BER | ATL | HND | BCN |
| – | TLV | DUB | FCO | RUH | SYD | FRU | BOG | SGN | ASU |
| – | CAI | BCN | AKL | BCN | SAW | AGP | MCO | ZQN | SJO |
| – | MAN | CPH | MNL | FCO | RUH | LIS | MAD | VLI | VVI |
| – | MIA | ZAG | JNB | DAR | MFM | BRU | HND | KIX | MXP |
| – | MXP | BER | MUC | LFW | AMS | SVO | DUB | TPE | UIO |
| – | BRU | CPT | GRU | LGW | HAN | ALC | MUC | WLG | LAX |
| – | CUN | LAX | DUB | DSS | LED | DYU | TPE | SEA | ZRH |
| – | NRT | BUD | CAI | KWI | PUS | MXP | ORD | CGK | EWR |
| – | MNL | MIA | MEX | DEL | BOM | CRL | FCO | SUV | IST |
| – | PMI | CRL | YVR | CPT | MEL | OSL | GRU | PVG | DXB |
| – | AUH | TLV | STN | KRT | AMM | OPO | LAS | OOL | GYE |
| – | LAX | BHX | SYD | LOS | DAC | LBD | DOH | FRA | DOH |
